# A Subset of Localized Prostate Cancer Displays an Immunogenic Phenotype Associated with Losses of Key Tumor Suppressor Genes

**DOI:** 10.1101/2021.01.27.21250464

**Authors:** Carla Calagua, Miriam Ficial, Caroline S. Jansen, Taghreed Hirz, Luke Del Balzo, Scott Wilkinson, Ross Lake, Anson T. Ku, Olga Voznesensky, David B. Sykes, Philip J. Saylor, Huihui Ye, Haydn Kissick, Sabina Signoretti, Adam G. Sowalsky, Steven P. Balk, David J. Einstein

## Abstract

**Purpose:** A subset of primary prostate cancer (PCa) expresses programmed death-ligand 1 (PD-L1), but whether they have unique tumor immune microenvironment (TIME) or genomic features is unclear.

**Experimental Design:** We selected PD-L1-positive high-grade and/or high-risk primary PCa, characterized tumor-infiltrating lymphocytes (TILS) with multiplex immunofluorescence, and identified genomic alterations in immunogenic and non-immunogenic tumor foci.

**Results:** One-quarter of aggressive localized PCa cases (29/115) had tumor PD-L1 expression >5%. This correlated with increased density of CD8^+^ T cells, a large fraction co-expressing PD-1, versus absent PD-1 expression on sparse CD8 T cells in unselected cases. Most CD8^+^PD-1^+^ cells did not express terminal exhaustion markers (TIM-3 or LAG-3), while a subset expressed TCF1. Consistent with these CD8^+^PD-1^+^TCF1^+^ cells being progenitors, they were found in antigen-presenting-cell niches in close proximity to MHC II^+^ cells. CD8 T cell density in immunogenic PCa and renal cell carcinoma (RCC) was nearly identical. Shallow *RB1* and *BRCA2* losses, and deep deletions of *CHD1*, were prevalent; the latter being strongly associated with a dendritic cell gene set in TCGA. Tumor mutation burden was variable; neither high microsatellite instability nor *CDK12* alterations were present.

**Conclusions:** A subset of localized PCa is immunogenic, manifested by PD-L1 expression and CD8^+^ T cell content comparable to RCC. The CD8^+^ T cells include effector cells and exhausted progenitor cells, which may be expanded by ICIs. Genomic losses of *RB1, BRCA2*, and *CHD1* may be drivers of this phenotype. These findings indicate that immunotherapies may be effective in biomarker-selected subpopulations of localized PCa patients.

**Statement of Translational Relevance:** Prostate cancer (PCa) is generally considered poorly immunogenic, with low expression of programmed death-ligand 1 (PD-L1) and low density of tumor-infiltrating immune cells. Accordingly, response rates to PD(L)-1 inhibition in unselected patients with advanced prostate cancer have been low. Here, we find that a substantial subset of aggressive primary PCa exhibits tumor PD-L1 expression and contains a high density of tumor-infiltrating lymphocytes. These lymphocytes contain sub-populations of exhausted progenitor CD8^+^ T cells and differentiated effector T cells, the hallmarks of ongoing anti-tumor immune response and a prerequisite for response to checkpoint inhibition. Furthermore, we identify genomic alterations that may be contributing to immunogenicity in these cases. These findings point to immune responses elicited in a subset of primary PCa, supporting the development of immune checkpoint blockade clinical trials in early-stage disease, such as biochemically recurrent PCa, that are driven by genomic features of the tumor or the immune microenvironment.

## Introduction

Immune checkpoint inhibitors (ICIs) that target the programmed cell death ligand-1 (PD-L1)/programmed cell death-1 (PD-1) pathway have demonstrated antitumor activity in a growing number of malignancies and are approved by the Food and Drug Administration for treatment of microsatellite instability-high (MSI-H)/mismatch repair-deficient (dMMR) or high tumor mutational burden (TMB) solid tumors. Broadly, tumor features associated with clinical response to anti-PD-1/PD-L1 therapy include T-cell infiltration, PD-L1 expression in tumor and/or immune cells, increased tumor mutational burden (TMB), and interferon gamma derived T-cell gene expression profiles (T-GEP) (1-4).

In advanced prostate cancer (PCa), the effect of anti-PD-1 ICI has been limited aside from the small subset of tumors that exhibit MSI-H/dMMR (5) or *CDK12* biallelic inactivation (6). Overall response rate (ORR) to pembrolizumab monotherapy has been estimated at 5-17% in unselected metastatic castration resistant prostate cancer (mCRPC) (7-9). Preliminary results of nivolumab plus ipilimumab (anti-CTLA4) in a mCRPC phase II trial (CHECKMATE-650) showed an ORR of 10% and 26% in patients with and without previous taxane-based chemotherapy, respectively (10). The impact of anti-PD-1 ICI in patients with non-metastatic disease remains unknown.

PD-L1 expression in PCa (≥5% of tumor cells staining positively) has been described from 15-52% in the primary setting (11-13) and from 4-50% in castration-resistant disease (14,15). Tumor heterogeneity, antibody selection, immunohistochemistry (IHC) protocols, and scoring are responsible for the variability of expression. To account for these differences, our group previously used two antibodies to evaluate PD-L1 expression in untreated radical prostatectomy (RP) specimens and matched RP cases that had been treated on trial with neoadjuvant hormonal therapy (medical castration plus abiraterone and prednisone) and had residual CRPC foci (16). This approach identified PD-L1 expression in 13.8% of localized PCa overall, and in 26.5% of Grade Group 4-5 (Gleason score 8-10) tumors, with no increase in neoadjuvant treated tumors. High numbers of CD8^+^ tumor infiltrating lymphocytes (TILs) positively correlated with PD-L1 expression. Interestingly, PD-L1 expression was more frequent in tumors from Black patients, consistent with data suggesting uniquely immunogenic phenotypes in this population (17-20).

These data suggest that a subset of localized PCa tumors, especially enriched among high-grade cases, are eliciting immune responses, which may be predictive of responsiveness to ICI. Both the expression of PD-L1 and increased number of TILs are reflective of interferon gamma exposure (21,22), and spatial distribution of TILs has identified three distinct cancer immune phenotypes: immune-excluded, immune-desert, and immune-inflamed (21). However, an inflamed microenvironment does not ensure response to ICI. Chronic inflammation, which is frequent in cancer, can result in an overactive immune response that ultimately leads to decreased cytotoxicity of CD8^+^ T cells (23-25). However, a subset of exhausted T-cells can be reinvigorated by ICI, leading to proliferation and activation of antitumor responses (26,27). Recently, transcription factor TCF-1 (encoded by *TCF7*) has been linked to T-cells with a “stem-like” phenotype, which have exhausted progenitor features and are associated with patient response to ICI (26-29). Other immune checkpoints associated with T cell exhaustion, like TIM-3, LAG-3, TIGIT, and VISTA, may also play a role in regulating anti-tumor T-cell response.

Here we selected a series of localized PCa cases based primarily on PD-L1 expression, with several additional cases having extensive TILs but low or absent PD-L1 expression, for genomic and microenvironment analyses. In contrast to the PD-1-negative antigen-naïve phenotype of sparse CD8^+^ T cells in unselected cases, the majority of CD8^+^ T cells in these selected cases had an effector phenotype, with a subset expressing TCF-1, indicative of exhausted progenitor cells that may respond to ICI. These were found within antigen-presenting cell (APC) niches. Genomic analysis showed significant enrichment for alterations in *RB1* and *BRCA2*, and deep deletions in *CHD1* were also prevalent. These results indicate that a subset of localized PCa (which we term immunogenic) are eliciting immune responses, that these tumors may have genomic features distinct from previously described drivers of immunogenicity, and may respond to ICI.

## Materials & Methods

### Tissue selection

In accordance with the U.S. Common Rule and after Institutional Review Board (IRB) approval, radical prostatectomy (RP) tissue (or biopsy cores alone in three cases) was collected retrospectively between 2007 and 2019 at Beth Israel Deaconess Medical Center (BIDMC, Boston, MA) and deidentified in accordance with BIDMC IRB protocol #2010-P-000254. Given previous findings of enriched PD-L1 expression and TILs among high-grade cases, we examined 115 total cases that were Gleason 8 or above (50 cases), or had other NCCN high-risk features (extracapsular extension or serum PSA >20 ng/dL; 54 cases). We identified 29 total cases (25% of screened cases) with tumor PD-L1 expression ≥5% by IHC; we included an additional three RP cases and three cases with only core biopsies available, all PD-L1-negative but with extremely high TILs based on histology, forming a cohort of 35 total cases (30% of screened cases). These cases comprised the “immunogenic” cohort. Additional MSI-H cases (also included in TCR sequencing analysis) were analyzed with multiplex panel #3 described below.

### Immunohistochemistry

Prostate tissue was fixed in formalin, processed, and embedded in paraffin using standard methods. For each case, the tissue block with the largest dimensions of the dominant tumor plus two or three additional blocks were selected for PD-L1 and CD3 IHC. Additional details are presented in **Supplemental Methods**.

### Immunogenicity criteria

Immunogenicity was defined as PD-L1 moderate to strong membranous staining in ≥5% of tumor cells, cytoplasmic staining was not considered. Extremely high density of TILs, defined as lymphocytes either present within the tumor cell nests or glands or immediately adjacent to tumor cells (i.e., distance between the lymphocyte and its nearest tumor cell being less than the diameter of an average tumor cell), was used to include six PD-L1-negative cases. All immunostains were evaluated by an experienced pathologist (H. Ye) and a trained MD investigator (C. Calagua).

### DNA isolation, hybrid capture, library preparation, and sequencing

We selected 11 immunogenic cases for DNA sequencing. We used Hematoxylin-Eosin and IHC (PD-L1, CD3) staining to annotate areas as immunogenic (PD-L1 ≥5% and extensive TILs), non-immunogenic (PD-L1 negative and no TILs), and normal (benign glands and/or stroma non-adjacent to tumor cells). We created tissue microarrays (TMAs) using the EZ Manual Microarray Kit (IHC WORLD). Each core measured 3 mm in diameter and TMAs included one or more immunogenic areas, one non-immunogenic area, and one benign area.

We extracted DNA, generated libraries, and performed hybrid capture with a custom bait panel targeting exons for 754 genes including those in commercial platforms, additional genes altered in PCa (30), and genes related to immune function. Full details are outlined in **Supplemental Methods.** We then performed targeted sequencing, with median on-bait coverage of 252× (range: 101× to 427×). We identified somatic and germline mutations, somatic copy number alterations, tumor mutational burden, and percent genome altered as outlined in the Supplement.

### Comparison with TCGA

GISTIC2-summarized copy number values were downloaded from cBioPortal for 489 prostate tumors from TCGA. Amongst cases where both *RB1* and *BRCA2* deletions were identified, the SEG file from each tumor was manually inspected using IGV to determine if the deletions were part of a single focal event (*i*.*e*. co-deleted) or occurred individually. A two-sided Fisher’s exact test was used to compare the frequency of focal co-occurrence events between the TCGA cohort and our immunogenic cohort. P values were adjusted (False Discovery Rate, FDR) using the method of Benjamini and Hochberg.

### TCR Sequencing

To investigate T-cell clonality, we extracted genomic DNA as above and sequenced the VDJ region of the T-cell receptor β chain using the immunoSEQ multiplex PCR-based method (Adaptive Biotechnologies, Seattle, WA). We used Adaptive Analyzer to analyze output.

### Multiplex immunofluorescence analysis

We used multiplex immunofluorescence (IF) panel 1 (see **Supplemental Methods** and **Supplemental Table 1**) to evaluate a set of unselected RP cases, spanning all grade groups. This seven-plex panel included antibodies to CD3, CD8, PD-1, FoxP3, CD68, CD163, along with DAPI counterstaining. Regions of interest included both immune-cell-rich and non-rich areas and included both tumor and benign areas. For analysis of CD8^+^ T cell density, we quantified lymphocytes present in gland plus stroma. For analysis of CD8 and PD-1 co-expression, we analyzed lymphocytes present in gland only (restricted to epithelial cells) since the regions of interest analyzed in comparison immunogenic cases were nearly devoid of stroma.

For multiplex IF panel 2 applied to immunogenic cases and to renal cell carcinoma cases, an in-house six-plex assay was optimized, including antibodies to CD8, PD-1, TIM-3, LAG-3, and TCF1, along with DAPI counterstain (**Supplemental Table 2**). Details of the image acquisition and image analysis workflow were reported previously (31).

Multiplex IF panel 3 consisted of a previously described panel of antibodies to CD8, PD-1, TCF1, and MHC II, and DAPI counterstaining (32) (**Supplemental Table 3**).

We performed comparisons of multiplex IF datasets with the Mann Whitney test using GraphPad Prism 9 (GraphPad Software, San Diego, CA). For multiple comparisons within multiplex IF datasets, we adjusted alpha using the Bonferroni correction; otherwise, alpha was set at 0.05.

## Results

### Subset of localized prostate cancer is PD-L1 positive and infiltrated with CD8^+^ effector cells

While immune infiltrates are generally uncommon in primary PCa, we observed previously that multiple tumor foci in 14-27% of localized PCa, enriched in high-grade cases, had PD-L1 expression and associated T-cell infiltrates (**Figure 1A-F**). To determine whether these cases may reflect a distinct immunogenic subset, we first examined tumor foci in unselected RP cases, spanning all Grade Groups, with multiplex IF panel 1 for CD8 and PD-1 expression. In these cases, as expected, CD8^+^ infiltrate was quite low, with a median density of 54 cells/mm^2^ (**Figure 1G)**. In addition, a very small percentage of the already rare CD8^+^ cells co-expressed PD-1, indicating they were primarily antigen-naïve (**Figure 1H, Supplemental Table 4**).

**Figure 1.**
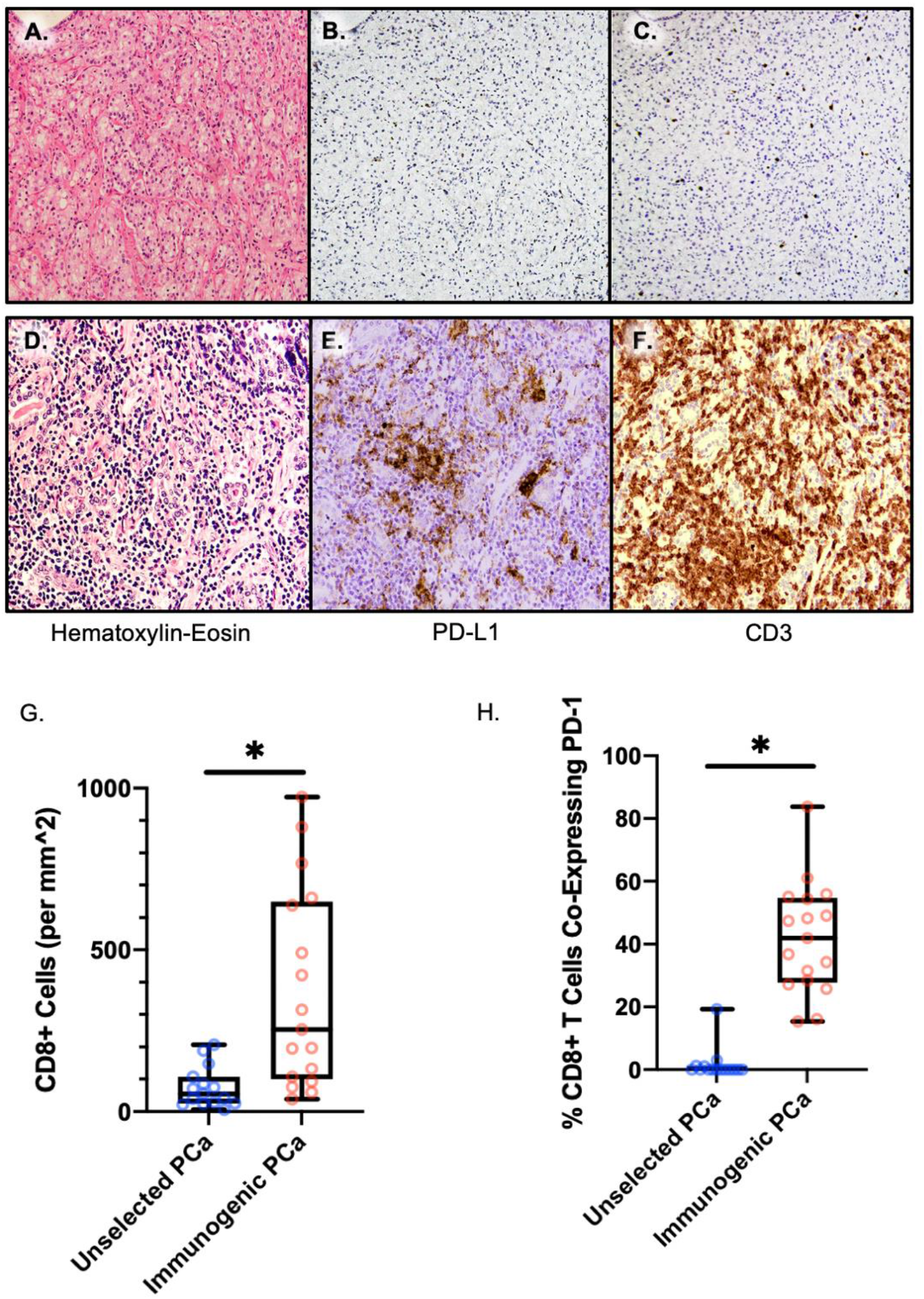
PD-1 expression by CD8^+^ T cells in unselected versus PD-L1 expressing cases. (A-F) Representative images of Hematoxylin-Eosin stain, PD-L1 IHC, and CD3 IHC in (A-C) non-immunogenic focus and (D-F) immunogenic focus within one immunogenic PCa case. (G, H) CD8^+^ tumor-infiltrating lymphocyte density (G), and co-expression of PD-1 among CD8^+^ cells (H), in unselected versus immunogenic prostate cancer.

We next examined a series of primary PCa cases selected primarily on the basis of PD-L1 expression, which we termed “immunogenic.” We characterized TILs from these cases using multiplex IF panel that included CD8 and PD-1 (IF panel 2). Tumor foci in these cases were highly enriched in CD8^+^ TILs compared with unselected prostate cancers (p=0.0002, **Figure 1G**). The median CD8^+^ cell density was 253 cells/mm2 (**Supplemental Table 5**). Furthermore, PD-1 expression indicated that these TILs were much more antigen-exposed: a median of 42% of CD8^+^ cells co-expressed PD-1 in this prostatectomy cohort, compared with a median of 0% in unselected prostatectomy cases (p<0.0001) (**Figure 1H**). Of note, the unselected cases were examined with a differently optimized IF panel and looked at random tumor foci, so may underestimate total CD8^+^ T cell content in cases where T-cell-infiltrated foci were present, but infrequent or rare. Nonetheless, these data indicate tumor foci with extensive CD8^+^ T cell infiltration are found in ∼30% of aggressive localized prostate cancer, and that these CD8^+^ T cells are qualitatively distinct in having evidence of antigen exposure.

### Tumor-infiltrating lymphocytes include exhausted progenitor and terminally differentiated populations

We further investigated CD8^+^ lymphocyte phenotypes in these TIL-infiltrated cases by multiplex IF with a panel against CD8, PD-1, LAG-3, TIM-3, and TCF1 (IF panel 2) (**Figure 2A**). The large majority (median 85%) of CD8^+^PD-1^+^ cells were negative for TIM-3 and LAG-3, indicating that they were not terminally exhausted, whereas 15% (median) co-expressed either TIM-3 or LAG-3, suggestive of terminal exhaustion (**Figure 2B**). Within the CD8^+^PD-1^+^ TIM-3(-) LAG-3(-) subset we found that a median of 34% co-expressed TCF1, suggestive of an exhausted progenitor phenotype, whereas a median of 66% did not co-express TCF1, TIM-3, or LAG-3, indicating possible differentiated effector status. As expected, TCF1^+^ cells rarely co-expressed TIM-3 or LAG-3 (**Supplemental Table 6)**. Overall, PD-1 was the most expressed exhaustion marker, followed by TIM-3.

**Figure 2.**
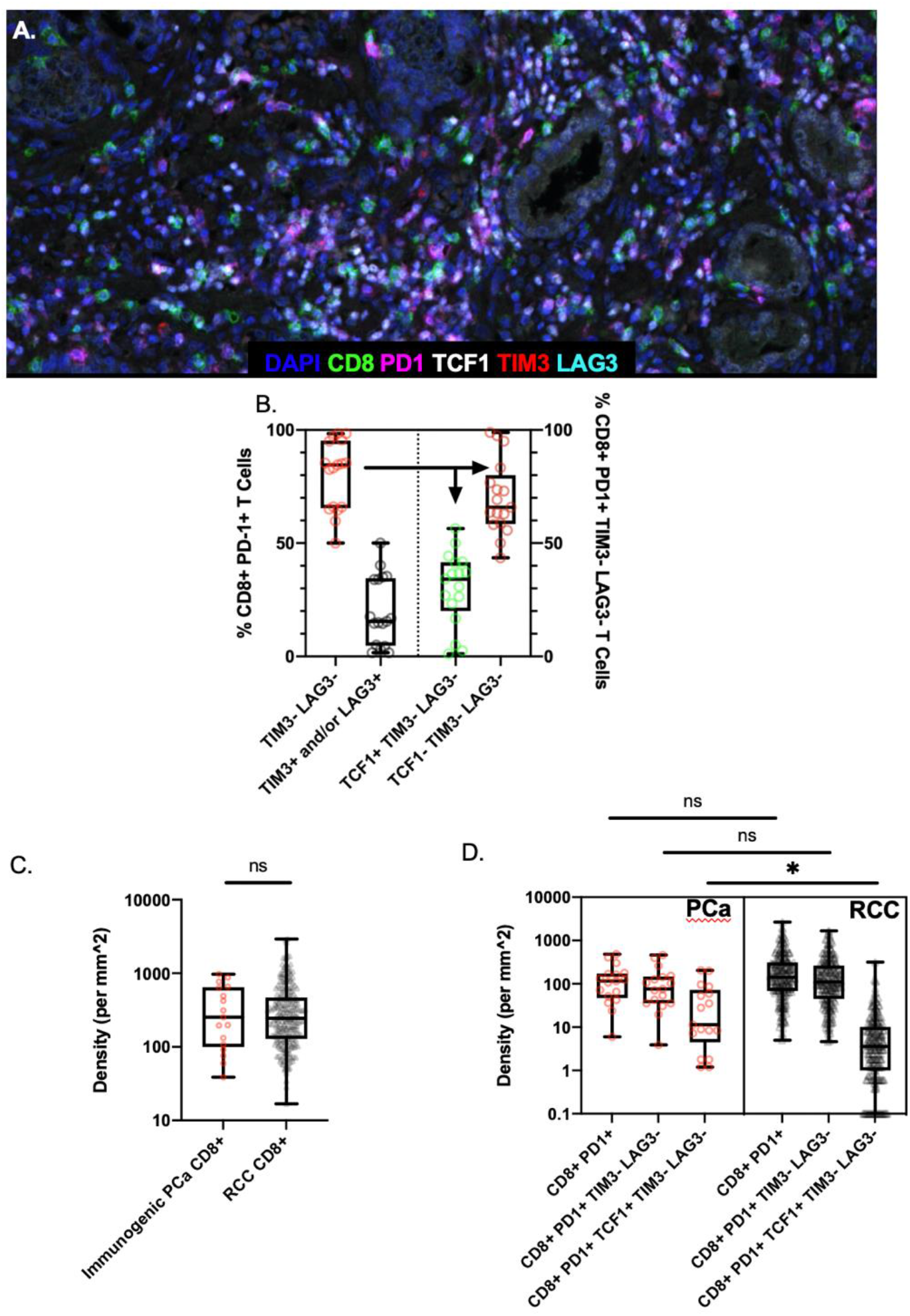
Multiplex immunofluorescence (IF) of CD8^+^PD-1^+^ immunogenic foci within immunogenic prostatectomy and RCC specimens. (A) A representative multiplex IF image in an immunogenic focus. (B) Left: median, interquartile ranges, and range of TIM-3 and/or LAG-3 co-expression in CD8^+^PD-1^+^ TILs. Right: median, interquartile ranges, and range of TCF1 co-expression in CD8^+^PD-1^+^ TIM-3(-) LAG-3(-) TILs. (C-D) Comparison of CD8^+^ TILs in immunogenic prostate cancer (red circles) versus metastatic renal cell carcinoma (grey triangles) in terms of CD8^+^ TIL density (C) and subsets of CD8^+^ TILs (D).

### Exhausted progenitor T cells are associated with antigen-presenting cell niches

In murine models of chronic viral infection, the T cell response is known to be maintained by a progenitor type cell that expresses the transcription factor TCF1 and resides predominantly in the lymphoid tissue (26). Interestingly, a number of studies reported the identification of analogous CD8^+^TCF1^+^ exhausted progenitor T lymphocytes directly in tumor tissue (26,29,33), and we previously observed these cells in close proximity to MHCII+ antigen-presenting cells (APCs). These aggregated TCF1^+^ T cells and MHC-II^+^ APCs form intra-tumoral APC niches (32), which closely resemble the T cell zone of lymphoid tissue, suggesting that these niches may support TCF1+ stem-like cell survival by analogous mechanisms to those in the lymphoid tissue. These previous studies examined these TCF1^+^ T cells and this pattern of immune organization in predominantly classically immunogenic tumor types, with a small subset of analysis in unselected prostate cancer patients.

Here, we identified intra-tumoral APC niches in immunogenic PCa (**Figure 3A-D**), supporting the notion that stem-like T cell-APC niches are a crucial factor in a patient’s ability to mount a strong anti-tumor immune response. As in prior work, APC niches are defined as areas within tumor tissue with MHC-II^+^ cells and CD8^+^TCF1^+^ T cells identified in the same 10,000μm^2^ area. These niches were present in the described immunogenic PCa cohorts, as well as in MSI-H PCa cases, and the percentage of tumor with these APC niches was not significantly different when comparing the immunogenic PCa cohorts to MSI-H cases (p=0.15, **Figure 3E**). This suggests that APC niches are an important feature of tumors with a strong immune response, regardless of the features used to identify immunogenic cases. The percentage of tumor tissue occupied by APC niches loosely correlated with amount of infiltrating MHC II+ cells (R^2^=0.18, p=0.065; **Figure 3F**) and more tightly correlated with the number of infiltrating CD8^+^ and TCF1^+^ TILs (R^2^=0.57, P=0.0001 for both comparisons; **Figure 3G-H**), further supporting the role of these APC niches in maintaining the tumor T cell response.

**Figure 3.**
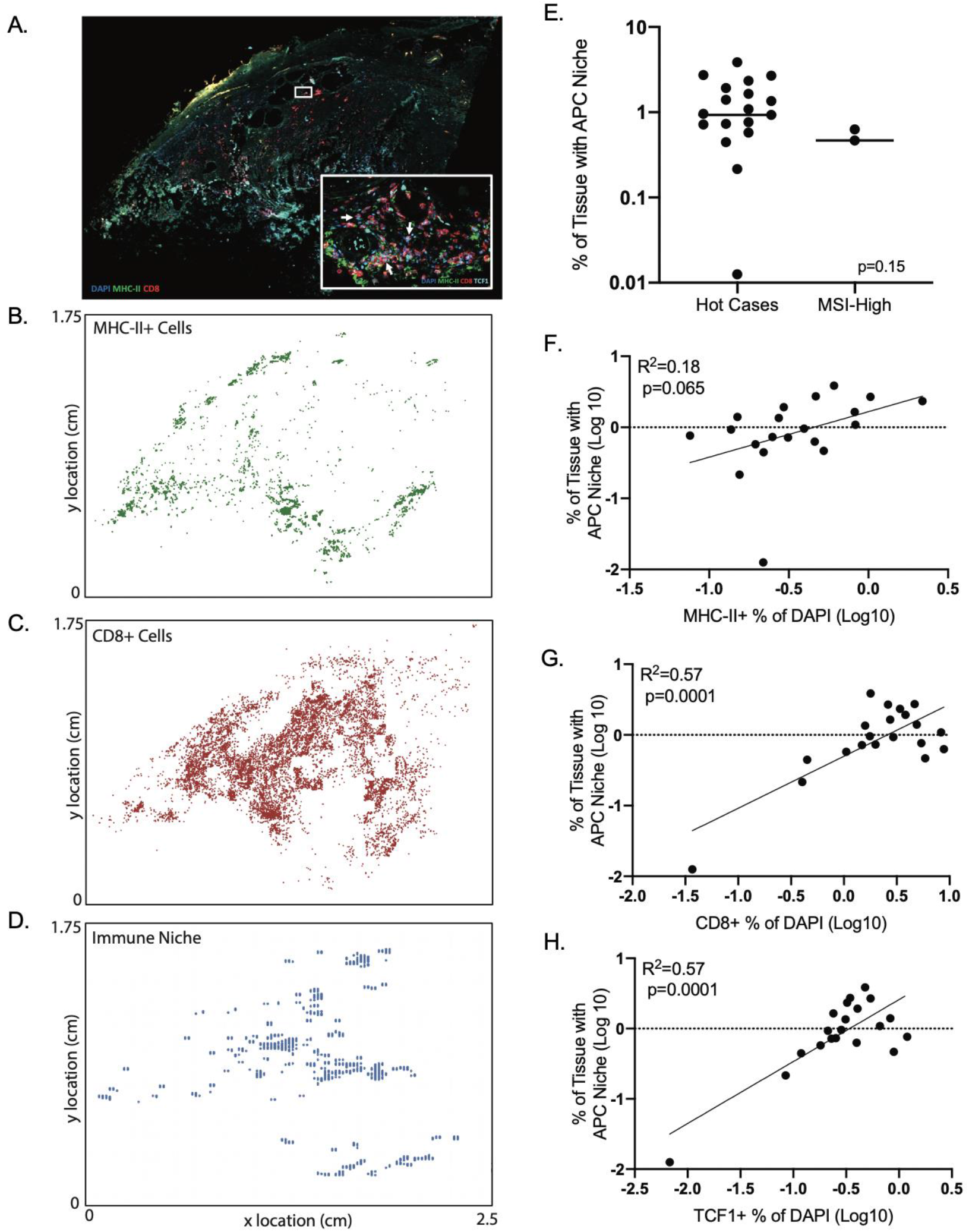
Evaluation of antigen-presenting cell (APC) niches within PCa cases. (A) Sample multiplex immunofluorescence image showing MHC II and CD8 expression. (B-D) Spatial analysis of individual cell populations and areas defined as APC immune niches (presence of both MHC-II+ and TCF1+ cells). (E) Percentage of tissue with APC niches by tissue cohort. (F-H) Correlation of percentage of tissue with APC niches with MHC-II+ Cells (F), CD8+ TILs (G), and TCF1^+^ TILs (H).

### Immune Infiltrates in Immunogenic Prostate Cancer are Comparable to those in Renal Cell Carcinoma

To benchmark our results, we compared them to results obtained with the same multiplex IF panel and same ROI selection in metastatic clear-cell renal cell carcinoma (mRCC) nephrectomy and metastatectomy specimens from the CheckMate-025 trial, a phase III randomized trial comparing nivolumab versus everolimus in patients with RCC previously treated with anti-angiogenic therapy. We recently characterized mRCC immune infiltrates in this trial (31). Median CD8^+^ density was nearly identical between immunogenic PCa and mRCC (p=1.00, **Figure 2C, Supplemental Table 7**). CD8^+^ TILs co-expressed PD-1 (and co-expressed PD-1 without TIM-3 or LAG-3) at comparable rates (p=0.22 and p=0.24, respectively). Interestingly, exhausted progenitors [CD8^+^PD-1^+^TCF1^+^ TIM-3(-) LAG-3(-)] were the one cell subtype seen at higher rates in immunogenic PCa compared with mRCC (p=0.0009, **Figure 2D**), which remained statistically significant with correction for multiple-comparison testing.

### Immunogenic localized prostate cancer demonstrates oligoclonal expansion of T cells

We sequenced the T-cell receptor β VDJ region in a subset of immunogenic cases, including a separate positive control case with MMR deficiency as demonstrated by loss of MSH2 and MSH6 by IHC (PCA25-MMR). Total productive rearrangements ranged from 2,480 to 13,645 (**Supplemental Figure 1**). Productive clonality ranged from 0.05 to 0.11. Maximum productive frequency ranged from 0.79% to 5.81%. The top clones were more expanded than less productive clones, suggesting oligoclonal expansion and tumor specificity. The top productive clones made up approximately 1-5% of total clones, and the top 10 productive clones made up approximately 5-15% of total. The MMR deficient case was comparable to the other cases along all parameters.

### Immunogenic localized prostate cancer is enriched for RB1, BRCA2, and CHD1 losses

We next carried out microdissection of tumor foci and exome sequencing in these immunogenic cases to identify genomic alterations. Of note, there was heterogeneity with respect to immune infiltrate in many cases, with some tumor foci near immunogenic foci having few immune cells and low or absent PD-L1 expression. Therefore, where possible we microdissected two or more immunogenic foci and a nonimmunogenic area from each case, as well as an area without tumor for germline DNA.

The most frequent alterations observed were in *BRCA2* and *RB1*, which are colocalized on chromosome 13 (**Figure 4, Supplemental Figure 2**). Shallow *RB1* deletions were found in immunogenic tumor foci from eight cases, and deep deletion was observed in one case (case 3, where a nonsense mutation was also found in one focus). In one additional case (Case 10) we found a shallow *RB1* deletion in the non-immunogenic focus, but not the immunogenic foci analyzed. This frequency of *RB1* alterations was higher (10 out of 11 cases) than in TCGA (218 out of 489) (p=0.003, P_Adj_=0.017) (**Figure 5A**). Based on these observations we carried out IHC for RB1 protein in a case where RB1 loss was found in immunogenic foci, but not in a non-immunogenic focus. This showed intact expression in the latter non-immunogenic focus and heterogeneous staining in the immunogenic foci (**Figure 5B**), suggesting that a subpopulation of tumor cells may have two-copy losses.

**Figure 4.**
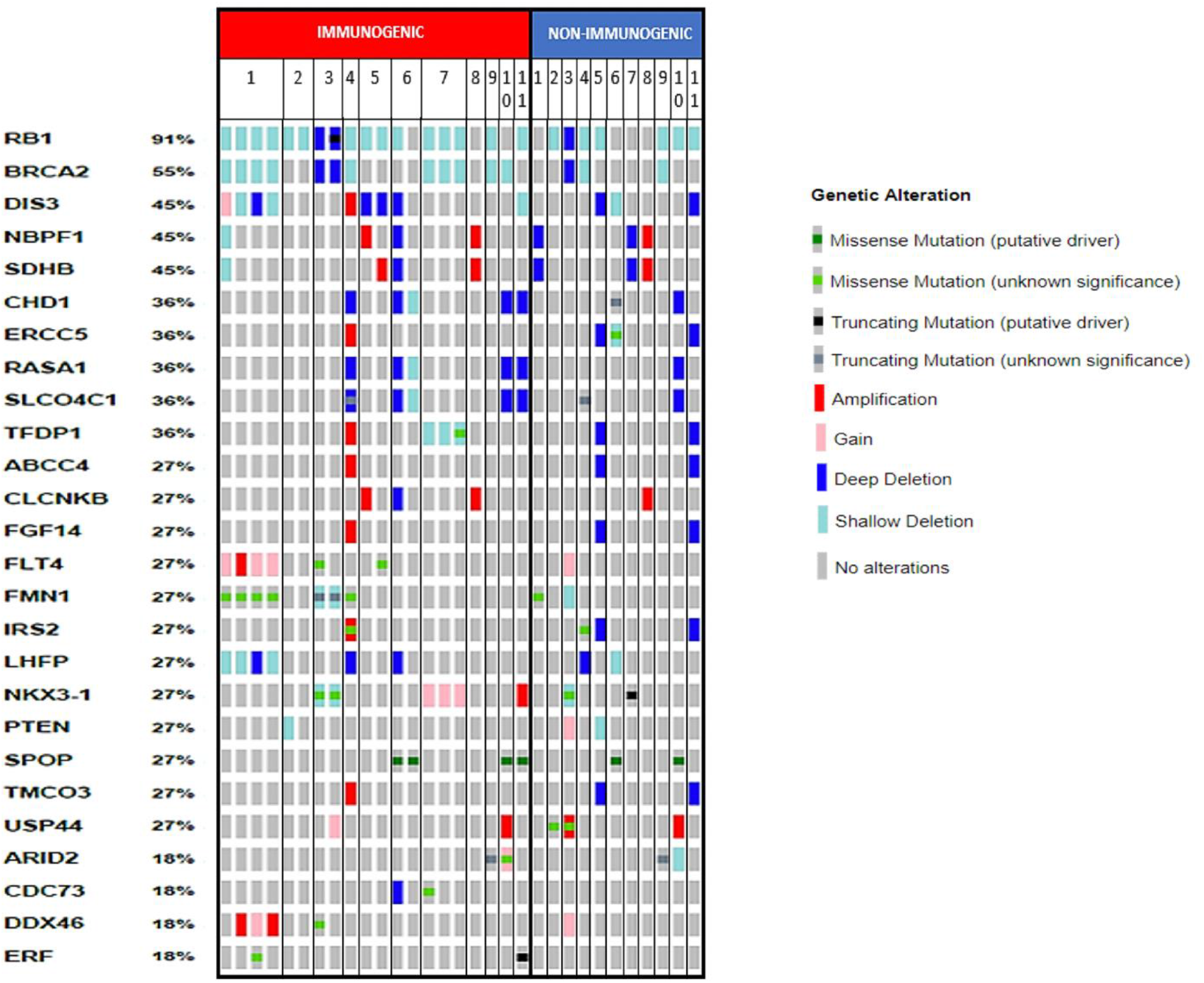
Genomic alterations in immunogenic tumors. Genomic alterations in immunogenic and non-immunogenic foci from immunogenic cases. Individual patients are designated by their numbers in the top row. Individual foci are represented by rectangles underneath their patient numbers. Percentages are based on number of cases with alterations in any focus divided by total number of cases.

**Figure 5.**
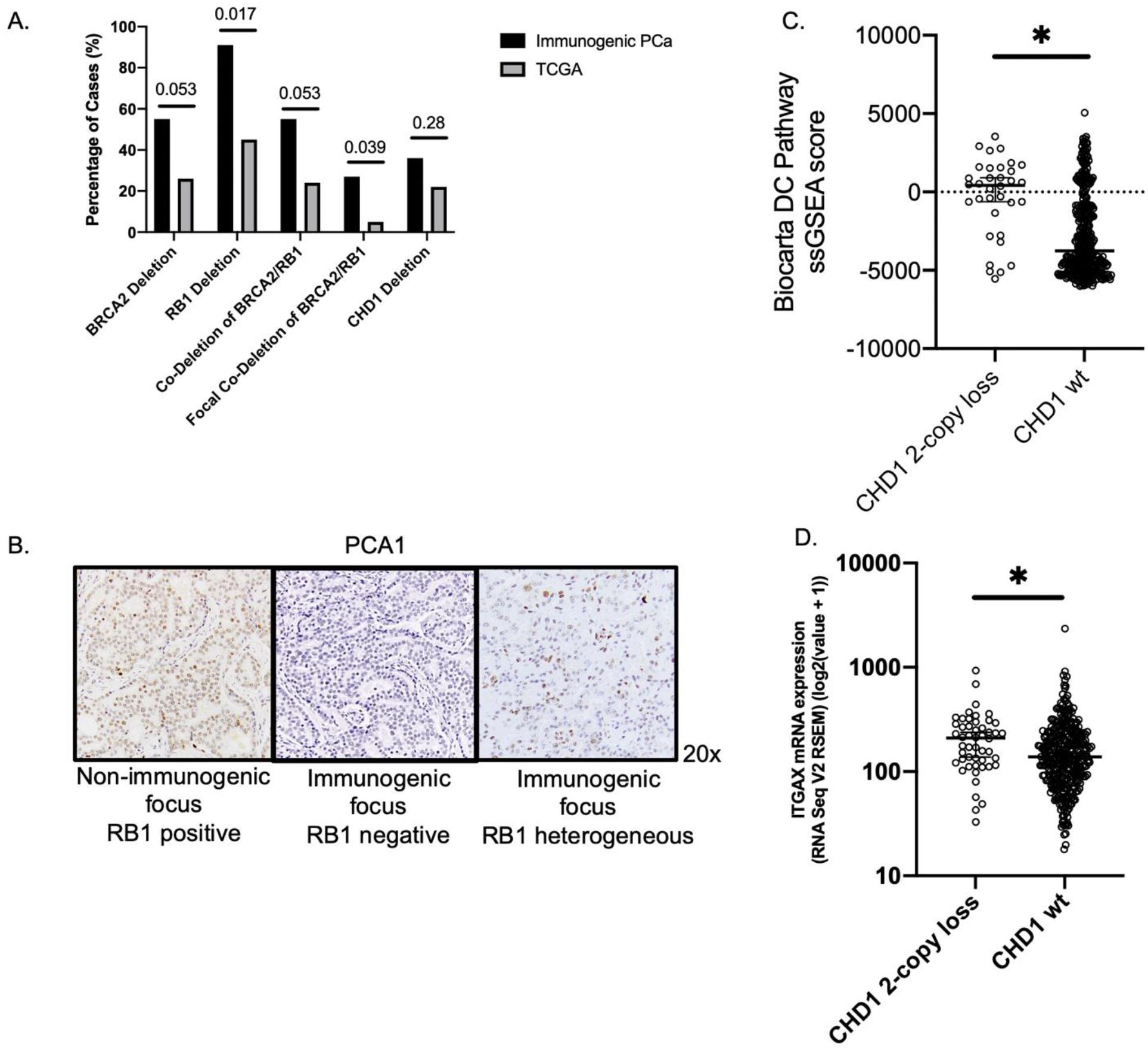
Loss of key tumor suppressor genes is associated with immunogenic phenotype. (A) Deletions in *BRCA2, RB1*, and *CHD1* in immunogenic PCa versus TCGA cases. P_Adj_ values are displayed above each comparison. (B) RB1 immunohistochemistry illustrating intact RB1 in a non-immunogenic focus, RB1 loss in an immunogenic focus, and heterogeneous RB1 loss in another immunogenic focus, all from one case (PCA1). (C) Single-sample Gene Set Enrichment Analysis of CHD1 2-copy loss versus wild-type (wt) TCGA cases illustrating difference in enrichment of the Biocarta Dendritic Cell (DC) Pathway geneset. (D) Expression of the dendritic cell marker *ITGAX* (CD11c) in TCGA cases with deep *CHD1* losses versus wild type (wt).

Five of these cases with *RB1* loss also had corresponding losses of *BRCA2*, which is located adjacent to *RB1* on chromosome 13. Of note, two of these cases with *BRCA2* loss also had germline frameshifts in *BRCA2* (Cases 1 and 3). The frequency of *BRCA2* loss (six out of 11) was higher than in TCGA (126 out of 489) (p=0.042, P_Adj_=0.053) (**Figure 5A**). Focal *BRCA2/RB1* co-deletion has recently been described as a marker of aggressiveness and possible sensitivity to poly-ADP-ribose polymerase inhibitors (34). In the immunogenic cases, the frequency of focal clonal co-deletion (3 out of 11) was significantly enriched compared to prostate TCGA cases (23 out of 489) (p=0.016, P_Adj_=0.039) (**Figure 5A**).

Amongst other genes recurrently altered and involved in DDR, we also found deep deletions in *CHD1* in 4 cases, which is greater than the ∼5-10% frequency of deep deletions found in unselected locally advanced or metastatic PCa (although statistical comparison with TCGA data did not reach statistical significance, likely due to small numbers and limited power) (**Figure 5A**). We then performed single-sample gene set enrichment analysis (ssGSEA), comparing TCGA tumors with deep CHD1 loss versus wild-type (wt). The top differentially expressed geneset was BIOCARTA_DC_PATHWAY (p<0.001, P_Adj_= 0.003) (**Figure 5C**). This appeared to be driven predominantly by increased expression of the dendritic cell marker *ITGAX* (CD11c) in CHD1-loss tumors (p=0.003, **Figure 5D**). Interestingly in all cases there were corresponding losses in *RASA1*, a suppressor of RAS activity. While this may reflect proximity on chromosome 5, the co-deletion of *CHD1* and *RASA1* is rare in unselected cases. Pathogenic *SPOP* mutations were also found in 3 of the cases with *CHD1* loss, consistent with a previously reported association between *CHD1* loss and *SPOP* mutations (35). In 2 cases we found deep deletions of *ERCC5* in non-immunogenic foci, but not the corresponding immunogenic foci.

Further potential tumor suppressor genes with mutations or losses in 3 or more cases were *DIS3, FMN1, LHFP*, and *SLCO4C1* (which may be linked to *CHD1*). A missense mutation was found in *TP53* in the immunogenic and non-immunogenic foci from one case. Interestingly, we did not observe deep losses in *PTEN*, which has been linked with immunosuppressive TIME (36), which would be expected in a substantial minority of aggressive PCa. The non-immunogenic foci analyzed in all cases had some shared alterations with the corresponding immunogenic foci, indicating they were clonal. However, in some cases there were additional alterations in the immunogenic foci. Several cases with shallow *RB1* and *BRCA2* losses in the immunogenic foci appeared to be wildtype in the corresponding non-immunogenic foci. Notably, three of four cases with deep *CHD1* loss in immunogenic foci were wildtype in the corresponding non-immunogenic foci. *FMN1*, with a role in adherens junction formation, had mutations in immunogenic foci from two cases that were not detected in the corresponding non-immunogenic foci. Full results are displayed in **Supplemental Figures 3-5**.

### Immunogenic localized prostate cancer shows high rates of genomic instability and variable tumor mutational burden

Percent of genome altered (PGA), defined as the percentage of the genome affected by copy number variation, has been described as a prognostic biomarker in patients undergoing RP. Patients with tumors having PGA greater than the median of 5.4% had at significantly higher risk of relapse (37). PGA in immunogenic and non-immunogenic tumor foci from immunogenic cases was at or above 5.4% in all but three cases (PCA4, PCA5, and PCA8) (**Figure 6A, C)**. The overall median PGA was much higher in immunogenic foci (11.2%) versus all foci (1.2%), although within individual cases, the correlation between immunogenic and non-immunogenic foci PGA was variable. This could also be due to contamination of the “non-immunogenic” foci by neighboring immunogenic foci.

**Figure 6.**
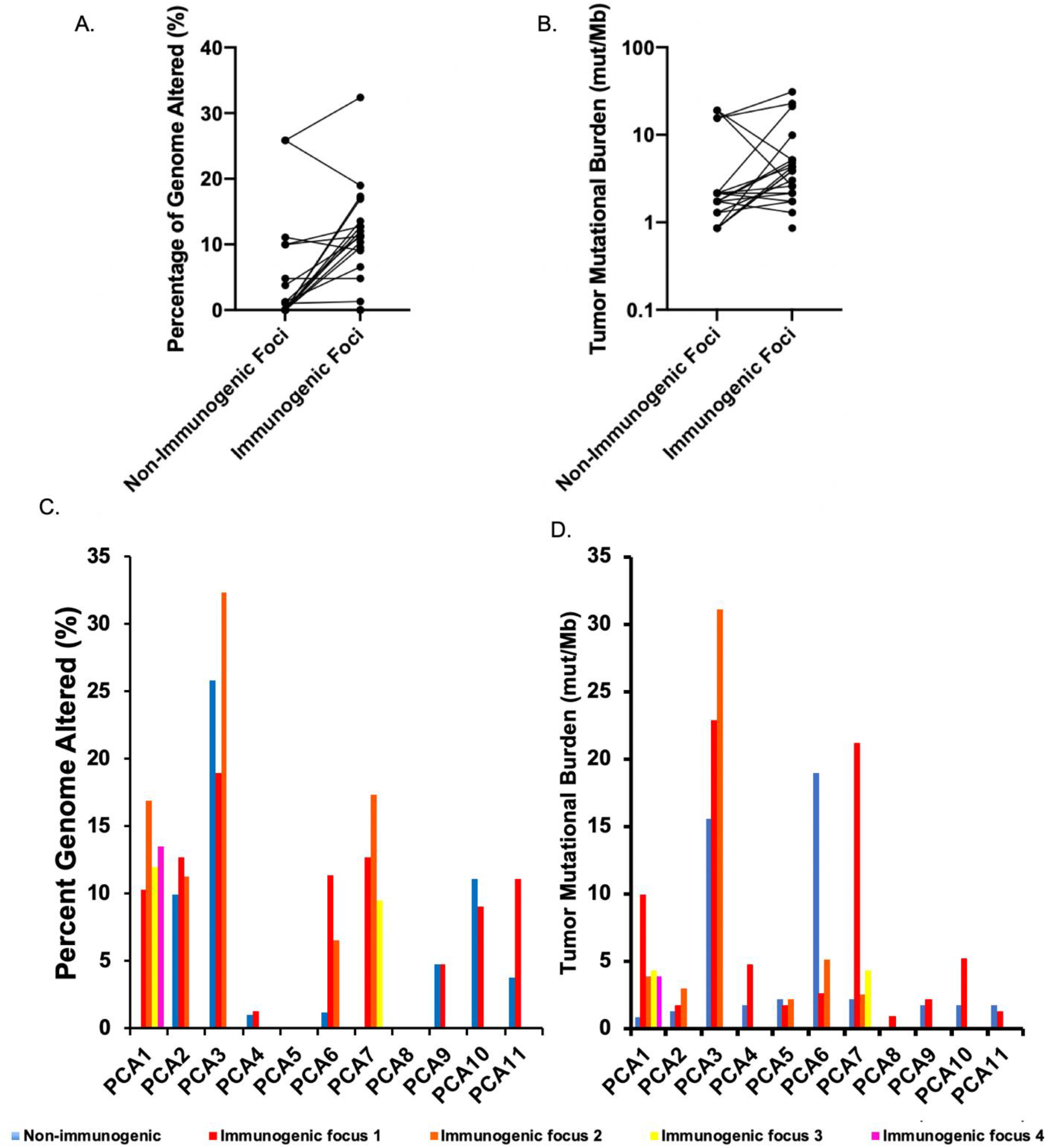
Genomic instability in immunogenic and non-immunogenic foci from immunogenic tumors. (A-D) Percentage of genome altered and tumor mutational burden in immunogenic and non-immunogenic foci from individual cases, shown as median of immunogenic foci (A-B) and as individual foci (C-D).

Tumor mutational burden (TMB), the number of non-synonymous mutations present per megabase (mut/Mb), is a potential biomarker for response to immunotherapy. Median TMB in these immunogenic cases was 1.73 mut/Mb in the TIL(-) PD-L1(-) non-immunogenic foci and 3.88 mut/Mb in TIL-high PD-L1^+^ immunogenic foci (**Figure 6B**). The latter is higher than the 1.36 mut/Mb and 2.93 mut/Mb previously described in primary and metastatic PCa, respectively (38), but lower than the threshold of 10 mut/Mb recently used for a tumor-agnostic FDA approval of PD-1 inhibition. TMB was not consistently higher in immunogenic foci compared to the corresponded non-immunogenic focus (**Figure 6B**). However, four cases did exhibit TMB of 10 mut/Mb or greater; notably, TMB was heterogeneous even across immunogenic foci within these cases, meaning that low TMB in a core biopsy of one area of the prostate would not rule out high TMB in another area (**Figure 6D)**. A small subset of advanced PCa may harbor mismatch repair deficiency leading to microsatellite instability and high TMB. In the immunogenic cases, we did not find mismatch repair alterations by next-generation sequencing or immunohistochemistry for the four mismatch repair proteins, nor did we identify any POLE mutations.

## Discussion

PCa has been considered to be a poorly immunogenic tumor, with low expression of PD-L1, sparse T cell infiltration, and little clinical benefit in response to ICI (7,39). In contrast to this general characterization, we demonstrate that a substantial subset of localized PCa, approximately one-quarter of the screened high-grade or high-risk cases, demonstrates tumor PD-L1 expression and substantial CD8^+^ T cell infiltration. The CD8^+^ infiltrates in these cases include exhausted progenitor (expressing TCF1) and effector phenotypes, and are comparable to those found in metastatic RCC, which is among the most immune-infiltrated cancer types.

TCF1 is necessary to support a progenitor CD8^+^ T cell population by repressing pro-exhaustion factors and inducing BCL6, a promoter of progenitor phenotype (40). It also appears to be essential for response to both vaccination and combined CTLA-4/PD-1 blockade in mouse models (26,29) and has been characterized as a predictor of response to ICI in melanoma (33). Exhausted progenitor cells are found in close proximity to APCs in localized prostate, kidney, and bladder specimens, and they give rise to clonally related terminally differentiated T cells via epigenetic de-repression of key genes involved in differentiation (32). Compared with metastatic RCC (mRCC), immunogenic PCa appears to contain a higher density of exhausted progenitors. In Checkmate-025, a trial comparing PD-1 blockade versus everolimus in mRCC, we recently found that density of exhausted progenitors had a significant interaction with treatment arm with respect to progression-free survival (31). Taken together, in immunogenic PCa cases, the findings of exhausted progenitors in APC niches, differentiated effector T cells, and oligoclonal expansion imply an active and tumor-specific immune response.

Terminally differentiated T cells are the primary target of ICIs in clinical use and in development. Here, we find that PD-1 is the primary exhaustion marker expressed in immunogenic localized PCa, with some co-expression of TIM-3. Similarly, in CheckMate-010, a study of PD-1 blockade in advanced RCC, we found that PD-1^+^ TIM-3(-) LAG-3(-) comprised approximately half of CD8^+^ TILs, with TIM3^+^ TILs present at lower levels and only rare LAG-3^+^ TILs (41). Percentage of CD8^+^PD-1^+^ TIM-3(-) LAG-3(-) was associated with improved clinical outcomes, potentially due to a subpopulation of TCF1^+^ cells, although this was not evaluated. Moreover, the combination of two biomarkers—high tumor PD-L1 expression and high percentage of CD8^+^PD-1^+^ TIM-3(-) LAG-3(-) TILs—identified a population of patients that had particularly good outcomes with PD-1 blockade. Conversely, absence of these two biomarkers was associated with a 0% response rate. We recently independently validated these findings in RCC in data from Checkmate-025 (31). Thus, our finding of a CD8^+^PD-1^+^ population without TIM-3 or LAG-3 in immunogenic, PD-L1-expressing PCa mirrors findings in the most PD-1-blockade-responsive subset of kidney cancer.

TIM-3 is inversely correlated with TCF1 (26), making these two markers useful to assess the balance between progenitors and terminally differentiated cells. TIM-3 is an important marker that in combination with PD-1 is associated with a particularly exhausted (but importantly, not senescent) T cell phenotype, and it can be co-targeted along with PD-1 for potentially synergistic effects (42,43). In localized kidney cancer, the percentage of CD8^+^PD-1^+^TIM-3^+^ TILs is the one CD8^+^ T cell subset that negatively correlates with clinical outcomes and has impaired function after stimulation (44). Similar findings were seen in ovarian high-grade serous carcinoma (45). TIM-3 represents a particularly attractive therapeutic target given its more restricted expression when compared with checkpoints such as CTLA-4, and thus potentially a more favorable toxicity profile.

We were able to identify a number of genomic alterations present in these immunogenic cases, and to differentiate genomic alterations between immunogenic and non-immunogenic foci within these cases. One key finding is the prevalence of alterations in DNA-damage repair genes, including *BRCA2* and *CHD1*, loss of which leads to decreased error-free homologous recombination repair of double-strand DNA breaks and therefore increased error-prone non-homologous-end-joining (NHEJ) repair (46). Another key finding is the marked prevalence of loss of *RB1*, primarily as shallow deletions, but with occasional deep deletions and one truncating mutation. Loss of *RB1* is associated with aggressive and castration-resistant prostate cancers, and it results in replicative stress. In particular, the co-alteration of *BRCA2* and *RB1*, even as single-copy loss, has been recently identified as associated with a uniquely aggressive subtype of localized prostate cancer (34). Pending confirmation in a larger cohort, the prevalence of these findings raises the hypothesis that these alterations result in replicative stress and more error prone NHEJ repair. This may result in increased mutational burden, but also in disrupted mitosis and cytoplasmic DNA, leading to Stimulator of Interferon Genes (STING) pathway activation.

Our analysis has so far primarily focused on infiltrating T cells, but the immune microenvironment clearly contains other elements, especially in advanced disease. Single-cell RNA sequencing of advanced PCa bone metastases has revealed depletion of B cells and an increase in M2-polarized tumor-associated macrophages (47). This is associated with increased exhaustion and decreased cytotoxicity programs in cytotoxic T lymphocytes in a CCR6-CCL20-dependent manner. Similarly, myeloid-derived suppressor cells (MDSCs) emerge in the context of castration resistance (48) and could be a mechanism of resistance to ICIs in advanced CRPC. Intriguingly, *CHD1* loss has been recently linked via IL-6 signaling with reduced MDSCs and increased CD8^+^ T cells in PCa models (49), suggesting one mechanism through which *CHD1* loss may drive immunogenicity. In our immunogenic tumors with deep *CHD1* deletions, *SPOP* inactivating mutations were commonly present. This association has been noted previously and linked to enhanced DNA damage, but in an immune context may also reflect SPOP activity as a ubiquitin ligase for PD-L1. Notably, deep *CHD1* deletions were highly associated with increased dendritic-cell-marker expression in TCGA, supporting an increase in APC activity in these tumors. Thus, *CHD1* loss, alone or in combinations with *SPOP* mutation, may drive immunogenicity by several mechanisms, and be a biomarker of response to ICI.

These data provide a rationale for testing ICI in earlier PCa spaces and with biomarker selection. We are currently accruing patients to a trial of PD-1 blockade in high-risk biochemically recurrent prostate cancer, over-sampling for PD-L1-expressing primary tumors (NCT03637543), and will test for correlations between the features identified above and clinical response. Another trial is testing PD-1 blockade in combination with chemotherapy for newly metastatic castration-sensitive prostate cancer, with genomic and proteomic biomarker selection (NCT04126070). Interestingly, while most patients with de novo metastatic PCa appear to have non-immunogenic tumors, there is a high-TIL subset, similarly correlated with high-grade histology (50), as in our localized tumors. Finally, further biomarker work may allow for the identification of immunogenic tumors prior to RP and subsequent development of biomarker driven neoadjuvant immunotherapy trials that exploit intratumoral progenitor T cell populations.

## Data Availability

Data available upon request

## Acknowledgements

The authors gratefully acknowledge the patients and the families of patients who contributed to this study. We wish to acknowledge Ying Huang, PhD and the Molecular Pathology Core Lab at Dana-Farber Cancer Institute for providing multiplex IF on unselected prostatectomy samples. Portions of this work utilized the computational resources of the NIH HPC Biowulf cluster.

## References

1. Cristescu R, Mogg R, Ayers M, Albright A, Murphy E, Yearley J, et al. Pan-tumor genomic biomarkers for PD-1 checkpoint blockade–based immunotherapy. Science 2018;362(6411).

2. Jerby-Arnon L, Shah P, Cuoco MS, Rodman C, Su M-J, Melms JC, et al. A cancer cell program promotes T cell exclusion and resistance to checkpoint blockade. Cell 2018;175(4):984–97. e24.

3. Thorsson V, Gibbs DL, Brown SD, Wolf D, Bortone DS, Yang T-HO, et al. The immune landscape of cancer. Immunity 2018;48(4):812–30. e14.

4. Lu S, Stein JE, Rimm DL, Wang DW, Bell JM, Johnson DB, et al. Comparison of biomarker modalities for predicting response to PD-1/PD-L1 checkpoint blockade: a systematic review and meta-analysis. JAMA oncology 2019;5(8):1195–204.

5. Abida W, Cheng ML, Armenia J, Middha S, Autio KA, Vargas HA, et al. Analysis of the prevalence of microsatellite instability in prostate cancer and response to immune checkpoint blockade. JAMA oncology 2019;5(4):471–8.

6. Wu Y-M, Cieślik M, Lonigro RJ, Vats P, Reimers MA, Cao X, et al. Inactivation of CDK12 delineates a distinct immunogenic class of advanced prostate cancer. Cell 2018;173(7):1770-82. e14.

7. Hansen A, Massard C, Ott P, Haas N, Lopez J, Ejadi S, et al. Pembrolizumab for advanced prostate adenocarcinoma: findings of the KEYNOTE-028 study. Annals of Oncology 2018;29(8):1807–13.

8. Antonarakis ES, Piulats JM, Gross-Goupil M, Goh J, Ojamaa K, Hoimes CJ, et al. Pembrolizumab for treatment-refractory metastatic castration-resistant prostate cancer: Multicohort, open-label phase II KEYNOTE-199 study. Journal of Clinical Oncology 2020;38(5):395.

9. Topalian SL, Hodi FS, Brahmer JR, Gettinger SN, Smith DC, McDermott DF, et al. Safety, activity, and immune correlates of anti–PD-1 antibody in cancer. New England Journal of Medicine 2012;366(26):2443–54.

10. Sharma P, Pachynski RK, Narayan V, Fléchon A, Gravis G, Galsky MD, et al. Nivolumab plus Ipilimumab for metastatic castration-resistant prostate cancer: preliminary analysis of patients in the CheckMate 650 Trial. Cancer cell 2020;38(4):489-99. e3.

11. Martin AM, Nirschl TR, Nirschl CJ, Francica BJ, Kochel CM, van Bokhoven A, et al. Paucity of PD-L1 expression in prostate cancer: innate and adaptive immune resistance. Prostate cancer and prostatic diseases 2015;18(4):325–32.

12. Gevensleben H, Dietrich D, Golletz C, Steiner S, Jung M, Thiesler T, et al. The immune checkpoint regulator PD-L1 is highly expressed in aggressive primary prostate cancer. Clinical Cancer Research 2016;22(8):1969–77.

13. Sharma M, Yang Z, Miyamoto H. Immunohistochemistry of immune checkpoint markers PD-1 and PD-L1 in prostate cancer. Medicine 2019;98(38).

14. Massari F, Ciccarese C, Calio A, Munari E, Cima L, Porcaro AB, et al. Magnitude of PD-1, PD-L1 and T lymphocyte expression on tissue from castration-resistant prostate adenocarcinoma: an exploratory analysis. Targeted oncology 2016;11(3):345–51.

15. Fankhauser CD, Schüffler PJ, Gillessen S, Omlin A, Rupp NJ, Rueschoff JH, et al. Comprehensive immunohistochemical analysis of PD-L1 shows scarce expression in castration-resistant prostate cancer. Oncotarget 2018;9(12):10284.

16. Calagua C, Russo J, Sun Y, Schaefer R, Lis R, Zhang Z, et al. Expression of PD-L1 in hormone-naïve and treated prostate cancer patients receiving neoadjuvant abiraterone acetate plus prednisone and leuprolide. Clinical Cancer Research 2017;23(22):6812–22.

17. Awasthi S, Berglund AE, Abraham-Miranda J, Rounbehler RJ, Kensler KH, Serna AN, et al. Comparative genomics reveals distinct immune-oncologic pathways in African American men with prostate cancer. Clinical Cancer Research 2020.

18. Yeyeodu ST, Kidd LR, Kimbro KS. Protective Innate Immune Variants in Racial/Ethnic Disparities of Breast and Prostate Cancer. Cancer immunology research 2019;7(9):1384–9.

19. Tang W, Wallace TA, Yi M, Magi-Galluzzi C, Dorsey TH, Onabajo OO, et al. IFNL4-ΔG allele is associated with an interferon signature in tumors and survival of African-American men with prostate cancer. Clinical Cancer Research 2018;24(21):5471–81.

20. Yuan J, Kensler KH, Hu Z, Zhang Y, Zhang T, Jiang J, et al. Integrative comparison of the genomic and transcriptomic landscape between prostate cancer patients of predominantly African or European genetic ancestry. PLoS Genetics 2020;16(2):e1008641.

21. Chen DS, Mellman I. Elements of cancer immunity and the cancer–immune set point. Nature 2017;541(7637):321–30.

22. Ayers M, Lunceford J, Nebozhyn M, Murphy E, Loboda A, Kaufman DR, et al. IFN-γ– related mRNA profile predicts clinical response to PD-1 blockade. The Journal of clinical investigation 2017;127(8):2930–40.

23. McLane LM, Abdel-Hakeem MS, Wherry EJ. CD8 T cell exhaustion during chronic viral infection and cancer. Annual review of immunology 2019;37:457–95.

24. Miller BC, Sen DR, Al Abosy R, Bi K, Virkud YV, LaFleur MW, et al. Subsets of exhausted CD8+ T cells differentially mediate tumor control and respond to checkpoint blockade. Nature immunology 2019;20(3):326–36.

25. Bengsch B, Ohtani T, Khan O, Setty M, Manne S, O’Brien S, et al. Epigenomic-guided mass cytometry profiling reveals disease-specific features of exhausted CD8 T cells. Immunity 2018;48(5):1029–45. e5.

26. Im SJ, Hashimoto M, Gerner MY, Lee J, Kissick HT, Burger MC, et al. Defining CD8+ T cells that provide the proliferative burst after PD-1 therapy. Nature 2016;537(7620):417–21.

27. Hudson WH, Gensheimer J, Hashimoto M, Wieland A, Valanparambil RM, Li P, et al. Proliferating transitory T cells with an effector-like transcriptional signature emerge from PD-1+ stem-like CD8+ T cells during chronic infection. Immunity 2019;51(6):1043-58. e4.

28. Chen Z, Ji Z, Ngiow SF, Manne S, Cai Z, Huang AC, et al. TCF-1-centered transcriptional network drives an effector versus exhausted CD8 T cell-fate decision. Immunity 2019;51(5):840-55. e5.

29. Siddiqui I, Schaeuble K, Chennupati V, Marraco SAF, Calderon-Copete S, Ferreira DP, et al. Intratumoral Tcf1+ PD-1+ CD8+ T cells with stem-like properties promote tumor control in response to vaccination and checkpoint blockade immunotherapy. Immunity 2019;50(1):195-211. e10.

30. Armenia J, Wankowicz SAM, Liu D, Gao J, Kundra R, Reznik E, et al. The long tail of oncogenic drivers in prostate cancer. Nat Genet 2018;50(5):645–51 doi 10.1038/s41588-018-0078-z.

31. Ficial M, Jegede OA, Sant’Angelo M, Hou Y, Flaifel A, Pignon JC, et al. Expression of T-cell Exhaustion Molecules and Human Endogenous Retroviruses as Predictive Biomarkers for response to Nivolumab in Metastatic Clear Cell Renal Cell Carcinoma. Clin Cancer Res 2020 doi 10.1158/1078-0432.Ccr-20-3084.

32. Jansen CS, Prokhnevska N, Master VA, Sanda MG, Carlisle JW, Bilen MA, et al. An intra-tumoral niche maintains and differentiates stem-like CD8 T cells. Nature 2019;576(7787):465–70.

33. Sade-Feldman M, Yizhak K, Bjorgaard SL, Ray JP, de Boer CG, Jenkins RW, et al. Defining T cell states associated with response to checkpoint immunotherapy in melanoma. Cell 2018;175(4):998-1013. e20.

34. Chakraborty G, Armenia J, Mazzu YZ, Nandakumar S, Stopsack KH, Atiq MO, et al. Significance of BRCA2 and RB1 co-loss in aggressive prostate cancer progression. Clinical Cancer Research 2020;26(8):2047–64.

35. Boysen G, Rodrigues DN, Rescigno P, Seed G, Dolling D, Riisnaes R, et al. SPOP-mutated/CHD1-deleted lethal prostate cancer and abiraterone sensitivity. Clinical cancer research 2018;24(22):5585–93.

36. Sharma MD, Shinde R, McGaha TL, Huang L, Holmgaard RB, Wolchok JD, et al. The PTEN pathway in Tregs is a critical driver of the suppressive tumor microenvironment. Science advances 2015;1(10):e1500845.

37. Hieronymus H, Schultz N, Gopalan A, Carver BS, Chang MT, Xiao Y, et al. Copy number alteration burden predicts prostate cancer relapse. Proceedings of the National Academy of Sciences 2014;111(30):11139–44.

38. Armenia J, Wankowicz SA, Liu D, Gao J, Kundra R, Reznik E, et al. The long tail of oncogenic drivers in prostate cancer. Nature genetics 2018;50(5):645–51.

39. De Bono JS, Goh JC, Ojamaa K, Piulats Rodriguez JM, Drake CG, Hoimes CJ, et al. KEYNOTE-199: Pembrolizumab (pembro) for docetaxel-refractory metastatic castration-resistant prostate cancer (mCRPC). American Society of Clinical Oncology; 2018.

40. Wu T, Ji Y, Moseman EA, Xu HC, Manglani M, Kirby M, et al. The TCF1-Bcl6 axis counteracts type I interferon to repress exhaustion and maintain T cell stemness. Science immunology 2016;1(6).

41. Pignon J-C, Jegede O, Shukla SA, Braun DA, Horak CE, Wind-Rotolo M, et al. irRECIST for the evaluation of candidate biomarkers of response to nivolumab in metastatic clear cell renal cell carcinoma: analysis of a phase II prospective clinical trial. Clinical Cancer Research 2019;25(7):2174–84.

42. Fourcade J, Sun Z, Benallaoua M, Guillaume P, Luescher IF, Sander C, et al. Upregulation of Tim-3 and PD-1 expression is associated with tumor antigen–specific CD8+ T cell dysfunction in melanoma patients. Journal of Experimental Medicine 2010;207(10):2175–86.

43. Sakuishi K, Apetoh L, Sullivan JM, Blazar BR, Kuchroo VK, Anderson AC. Targeting Tim-3 and PD-1 pathways to reverse T cell exhaustion and restore anti-tumor immunity. Journal of Experimental Medicine 2010;207(10):2187–94.

44. Granier C, Dariane C, Combe P, Verkarre V, Urien S, Badoual C, et al. Tim-3 expression on tumor-infiltrating PD-1+ CD8+ T cells correlates with poor clinical outcome in renal cell carcinoma. Cancer research 2017;77(5):1075–82.

45. Fucikova J, Rakova J, Hensler M, Kasikova L, Belicova L, Hladikova K, et al. TIM-3 dictates functional orientation of the immune infiltrate in ovarian cancer. Clinical Cancer Research 2019;25(15):4820–31.

46. Shenoy T, Boysen G, Wang M, Xu Q, Guo W, Koh F, et al. CHD1 loss sensitizes prostate cancer to DNA damaging therapy by promoting error-prone double-strand break repair. Annals of Oncology 2017;28(7):1495–507.

47. Baryawno N, Kfoury Y, Severe N, Mei S, Gustafsson KU, Hirz T, et al. Human prostate cancer bone metastases have an actionable immunosuppressive microenvironment. bioRxiv 2020.

48. Calcinotto A, Spataro C, Zagato E, Di Mitri D, Gil V, Crespo M, et al. IL-23 secreted by myeloid cells drives castration-resistant prostate cancer. Nature 2018;559(7714):363–9.

49. Zhao D, Cai L, Lu X, Liang X, Li J, Chen P, et al. Chromatin Regulator CHD1 Remodels the Immunosuppressive Tumor Microenvironment in PTEN-Deficient Prostate Cancer. Cancer Discov 2020;10(9):1374-87 doi 10.1158/2159-8290.Cd-19-1352.

50. Mendes L GE, Parry M, Lall S, Santos Vidal S, Bautista C, Zakka L, Berney D, Attard G, James N. Tumour Infiltrating Lymphocytes assessment in the STAMPEDE cohort: association of brisk inflammation with high grade histology. EACR-AACR-ASPIC Tumor Microenvironment 2020. Lisbon, Portugal 2020.

