## Supplement for "A Subset of Localized Prostate Cancer Displays an Immunogenic Phenotype Associated with Losses of Key Tumor Suppressor Genes"

**Supplemental Methods**

*Immunohistochemistry*

The following antibodies were used for IHC staining: mouse monoclonal anti-PD-L1 antibody (E1L3N, Cell Signaling Technology; 1:200) and mouse polyclonal anti-CD3 antibody (Agilent, 1:400). Five-micron sections were baked at 60°C for an hour, followed by deparaffinization, rehydration, and epitope retrieval using the Dako PT Link platform (Agilent). IHC staining was carried out on the Dako Link 48 autostainer (Agilent), with antibody incubation for 60 minutes, amplification using Envision FLEX mouse linkers, and visualization using the Envision Flex High-sensitivity visualization system (Agilent).

*DNA isolation, hybrid capture, library preparation, and sequencing*

We cut TMAs into 20-micron sections and scraped each core with a blade into a 1.5 low adhesion microcentrifuge tube for DNA extraction using the Qiamp DNA FFPE Tissue Kit Cat# 56404 following the manufacture’s protocol. DNA concentration was measured by QUBIT DSDNA HS ASSAY (Thermo Fisher Cat# Q32854).

Agilent SureSelect XT HS part# G9704E was used to generate the libraries from 42 “hot”, “cold” and benign samples following the manufacturer’s protocol. 22 to 70 ng of input gDNA was used per sample. We performed 13 cycles of pre-hybridization PCR for all samples to amplify, index and barcode individual libraries prior to hybrid capture. Two to three libraries (500 ng of amplified DNA per library) were grouped into 18 pools corresponding to 11 patients, for pooled capture reactions. Hybrid capture was performed using a custom bait design that covers the coding regions of 800 cancer-related genes plus the entire locus for a subset of tumor suppressors and oncogenes (Design ID: 3183291; 56762 probes; 3.9Mb capture size). We performed 10 cycles of PCR to amplify post-capture material. QC was performed prior and post hybrid capture using the Agilent TapeStation platform.

Post-capture libraries were combined in equimolar ratios into two separate pools based on their unique library indices. The two pools were sequenced on an Illumina NovaSeq SP FlowCell with an XP2 loading kit, with 150 cycles paired-end (2x150). Pass-filter reads were demultiplexed into FASTQ files, trimmed using Agilent SureCallTrimmer version 4.0.1 and aligned at the lane-level to the human genome (version b37+decoy) with BWA-mem version 0.7.17. SAM files were sorted, duplicate-marked and base quality score recalibrated using the Genome Analysis Toolkit (GATK) version 4.1.3.0. Lane-level BAM files were merged using Picard and duplicate-marked again using GATK. Capture efficiency and coverage were determined using GATK CollectHSMetrics.

Somatic mutations were identified using GATK MuTect2, first by calling all normal samples in tumor-only mode to generate a panel-of-normals, and then running MuTect2 again in tumor-normal (paired) mode also against the panel-of-normals. VCF files were filtered by MuTect2 to remove false-positives, artifacts, and orientation-bias mutations. Germline mutations were identified using GATK HaplotypeCaller in cohort mode, combining and genotyping each VCF file using GATK CombineGVCFs and GenotypeGVCFs, respectively. The unfiltered VCF file was recalibrated for point mutations and insertions/deletions using VariantRecalibrator against HapMap 3.3, 1000 Genomes Omni 2.5, 1000 Genomes Phase 1 SNPs, dbSNP version 138, and the Mills/1000 Genomes gold-standard indel set. Filtered somatic and germline VCF files were annotated with Oncotator (version 1, April052016 data corpus). All mutations were individually inspected and curated using the Integrated Genome Viewer (IGV). Tumor mutation burden (TMB) was calculated by determining the number of point mutations at 5% or greater variant allele fraction from all coding regions of the genome covered by the capture library and dividing it by the number of bases of the coding regions of the genome covered by the capture library.

Somatic copy number alterations (SCNA) were inferred from the panel-based sequencing data using the GATK SCNA pipeline. First, read counts were determined from each tumor or normal BAM file using GATK CollectReadCounts, and a panel-of-normals was used to denoise the read counts of all tumor samples using GATK CreateReadCountPanelOfNormals and GATK DenoiseReadCounts, respectively. Next, b-allele frequencies were determined for all tumor and normal BAM files using GATK CollectAllelicCounts. Finally, GATK ModelSegments was used to combine b-allele data and smoothed read count for each tumor/normal pair to generated segmentation (SEG) files with whole-genome copy number estimates. Copy numbers were called using GATK CallCopyRatioSegments and modeled with regions of heterozygosity with GATK PlotModeledSegments. The final modeled SEG file was then processed using GISTIC 2.0 (GenePattern module version 6.15.28) to call gene-level gains or losses (threshold 0.1 for single-copy, 1.3 for two-copy). All gene-level copy number calls were manually inspected and curated using IGV. Percent genome altered (PGA) was calculated by adding the lengths of all genomic segments estimated to have greater than 0.1 or less than -0.1 log_2_ copy number ratios, and dividing it by the length of the human genome.

FASTQ files from each of the 489 prostate cancer TCGA cases were downloaded from the NCI Genomic Data Commons, aligned with STAR version 2.7.0f and gene expression summaries were estimated using RSEM. Transcripts per million (TPM) values for each gene were processed using single-sample geneset enrichment analysis against the mSigDB Oncogenic Gene Signatures genesets. A two-sided t-test was used to compare the average ssGSEA score for each geneset in the CHD1-deletion or CHD1-WT group, and then P values were adjusted using the method of Benjamini and Hochberg.

*Multiplex immunofluorescence (IF) analysis*

IF panel 1: A seven-plex IF assay was performed on 4-µm FFPE sections, using Leica Bond Rx autostainer. Briefly the staining consists of sequential tyramine signal amplified immunofluorescence labels for each target, and a DAPI counterstain. Each labeling cycle consists of application of a primary antibody, a secondary antibody conjugated to horse radish peroxidase (HRP), and an opal fluorophore (Opal 690, Opal 570, Opal 540, Opal 620, Opal 650 and Opal 520, Akoya Biosciences), respectively. The stained slides were scanned on a Perkin Elmer Vectra 3 imaging system (Akoya Biosciences) and analyzed using Halo Image Analysis platform (Indica Labs). Each single stained control slide is imaged with the established exposure time for creating the spectral library. We ran an algorithm learning tool utilizing the Halo image software training for the gland and stroma regions, and subsequently completed cell segmentation. The thresholds for the antibodies were set respectively, based on the staining intensity, by cross reviewing more than 20 images. Cells with the intensity above the setting threshold were defined as positive.

IF panel 2: The assay was performed on a Bond RX Autostainer (Leica Biosystems) using the Opal multiplex IHC system (PerkinElmer/Akoya Biosciences Cat# NEL871001KT), as described in detail previously (*31*) (DOI: [dx.doi.org/10.17504/protocols.io.bjbzkip6](https://dx.doi.org/10.17504/protocols.io.bjbzkip6)). Whole slide multispectral images were acquired at 10x magnification using Vectra 3.0 (PerkinElmer/Akoya Biosciences) and visually inspected by using Phenochart 1.0.9 (PerkinElmer/Akoya Biosciences, RRID:SCR_019160) to select five intra-tumoral CD8^+^ T cell-enriched regions of interest (ROIs; area of each ROI = 669 µm x 500 µm) per case. In one case, additional ROIs were taken to ensure that a minimum of 100 CD8^+^ cells would be analyzed. ROIs were scanned at 20x magnification, imported into InForm v2.2.0 (PerkinElmer, RRID:SCR_019161) and deconvoluted using a multispectral library built with single stain slides. Image analysis was performed using HALO v2.1.1637.18 (Indica Labs, RRID:SCR_018350) and the Indica Labs High-Plex FL v2.0 module. A unique algorithm was created for each case, and its accuracy validated through visual inspection by a pathologist (M. Ficial) with extensive experience in image analysis.

IF panel 3: We deparaffinized sections in successive incubations with xylene and decreasing concentrations (100, 95, 75, 50, 0%) of EtOH. Antigen retrieval utilized Abcam 100x TrisEDTA Antigen Retrieval Buffer (pH = 9) heated under high pressure. We washed sections in PBS + 0.1% Tween20 before antibody staining. We blocked sections for 30 min with a solution of 10% goat serum in 1x PBS + 0.1% Tween20. before staining with primary and secondary antibodies. Primary antibodies were used at a concentration of 1:100 (MHC-II) or 1:150 (CD8, TCF1) and incubated for 1 h at room temperature. Secondary antibodies were used at a concentration of 1:250 (A488, A568) or 1:500 (A647) and incubated for 30 min at room temperature. We collected IF images using a Zeiss Z.1 Slide Scanner equipped with a Colibri 7 Flexible Light Source. We used Zeiss ZenBlue software for post-acquisition image processing. We analysed images using CellProfiler and custom R and python scripts, as previously described (32). CellProfiler was used to identify cellular objects within images and measure fluorescence intensity. Custom R and python scripts were used to perform quality control and remove imaging artifacts, measure distance between cellular objects, and calculate cellular density.

**Supplemental Table 1:** Immunofluorescence Antibodies for Panel #1

| **Target** | **Antibody Type** | **Clone** | **Provider** |
| --- | --- | --- | --- |
| CD3 | Rabbit polyclonal | Polyclonal | Dako |
| CD8 | Mouse monoclonal | C8/144B | Dako |
| PD-1 | Mouse monoclonal | EH33 | Cell Signalling |
| FoxP3 | Rabbit monoclonal | D2W8E | Cell Signalling |
| CD68 | Mouse monoclonal | PG-M1 | Dako |
| CD163 | Mouse monoclonal | 10D6 | Leica Biosystem |

**Supplemental Table 2:** Immunofluorescence Antibodies for Panel #2

| **Primary Antibodies** | **Clone or reference #** | **Provider** | **Order of serially stained markers** | **Dilution** | **Link (dilution, incubation time, ref #, company)** | **Secondary (tertiary) HRP-conjugated antibody** | **Tyramide conjugated fluorochrome** |
| --- | --- | --- | --- | --- | --- | --- | --- |
| Goat anti-TIM-3 | AF2365 | R&D systems  (Cat# AF2365, RRID:AB_355235) | 1 | 1/1000 | rabbit-anti-goat (1/750, 15 minutes, E0466, Agilent) | BOND Polymer Refine Detection Kit (Leica Biosystems) | Opal 540 (1:50) |
| Mouse anti-LAG-3 | 17B4 | LifeSpan Biosciences  (Cat# LS‑B2237-50, RRID:AB_1650048) | 2 | 1/1000 | none | BOND Polymer Refine Detection Kit (Leica Biosystems) | Opal 620 (1:150) |
| Mouse anti-CD8 | C8/144B | Agilent  (Cat#  M710301-2, RRID:AB_2075537) | 3 | 1/5000 | none | BOND Polymer Refine Detection Kit (Leica Biosystems) | Opal 520 (1:150) |
| Mouse anti-PD-1 | EH33 | Dr. Gordon Freeman | 4 | 1/5000 | none | BOND Polymer Refine Detection Kit (Leica Biosystems) | Opal 690 (1:50) |
| Rabbit anti TCF7 | C63D9 | Cell Signaling  (Cat#2203S, RRID:AB_2199302) | 5 | 1/2000 | none | BOND Polymer Refine Detection Kit (Leica Biosystems) | Opal 570 (1:100) |

**Supplemental Table 3:** Immunofluorescence Antibodies for Panel #3

| **Target** | **Antibody Type** | **Clone** | **Concentration** | **Secondary** | **Concentration** |
| --- | --- | --- | --- | --- | --- |
| MHC-II (HLA-DR, DP, DQ) | Mouse IgG2a | Tu39 | 1:100 | Goat anti-mouse IgG2a A488 | 1:250 |
| TCF1 | Rabbit | C63D9 | 1:150 | Goat anti-rabbit A568 | 1:250 |
| CD8 | Mouse IgG1 | C8/144B | 1:150 | Goat anti-mouse IgG1 A647 | 1:500 |

**Supplemental Table 4:** Summary of CD8+ and CD8+PD-1+ infiltrates in unselected primary prostate cancer

|  | Gland-only Tumor CD8+ | Gland-only Benign CD8+ | Gland-only Tumor CD8+PD-1+ | Gland-only Benign CD8+PD-1+ |
| --- | --- | --- | --- | --- |
| Median Raw Count | 33 | 71 | 0 | 1 |
| Median % of Total Epithelial Cells | 1% | 2% | 0% | 0% |
| Median % of CD8+ Cells |  |  | 0% | 3% |
|  | Gland + Stroma Tumor CD8+ | Gland + Stroma Benign CD8+ | Gland + Stroma Tumor CD8+PD-1+ | Gland + Stroma Benign CD8+PD-1+ |
| Median Raw Count | 123 | 170 | 1 | 6 |
| Median % of Total Cells | 1% | 2% | 0% | 0% |
| Median % of CD8+ Cells |  |  | 1% | 3% |

**Supplemental Table 5:** Subsets of Tumor-Infiltrating Lymphocytes in Immunogenic Prostate Cancer Cases

|  | Percentage of Total DAPI+ Cells  *Median*  *(Range)* | Percentage of CD8+ Cells *Median (Range)* | Density  (cells/mm2)  *Median*  *(Range)* |
| --- | --- | --- | --- |
| CD8+ | 3% (0.1 - 10%) |  | 253  (39 - 973) |
| & PD-1+ |  | 42%  (15 - 84%) | 116 (6 - 484) |
| & TCF1+ |  | 19%  (0.6 - 41%) | 27 (1 - 397) |
| & TIM-3+ |  | 9%  (1 - 36%) | 27 (2 - 103) |
| & LAG-3+ |  | 2%  (0.5 - 18%) | 8 (0 - 51) |
| Antigen-exposed but not exhausted | | | |
| CD8+ PD-1+  TIM-3(-) LAG-3(-) |  | 29%  (10 - 60%) | 76 (3 - 460) |
| Terminally exhausted | | | |
| CD8+ PD-1+  TIM-3+ and/or LAG-3+ |  | 5  (0.0 - 30%) | 21 (1 - 58) |
| Stem-like progenitor | | | |
| CD8+ PD-1+ TCF1+  TIM-3(-) LAG-3(-) |  | 9% (0.6 - 23%) | 11 (1 - 204) |
| Differentiated effector, not terminally exhausted | | | |
| CD8+ PD-1+ TCF1(-)  TIM-3(-) LAG-3(-) |  | 21%  (7 - 57%) | 667  (3 - 256) |

**Supplemental Table 6:** TCF1 is rarely co-expressed with TIM-3 and/or LAG-3

|  | Percentage of CD8+ Cells *Median (Range)* |
| --- | --- |
| CD8+ PD-1+ TCF1+ TIM-3+ LAG-3(-) | 0.4% (0.0 - 3%) |
| CD8+ PD-1+ TCF1+ TIM-3(-) LAG-3+ | 0.0% (0.0 - 4%) |
| CD8+ PD-1+ TCF1+ TIM-3+ LAG-3+ | 0.0% (0.0 - 2%) |
| CD8+ PD-1(-) TCF1+ TIM-3+ LAG-3(-) | 0.3% (0 - 2%) |
| CD8+ PD-1(-) TCF1+ TIM-3(-) LAG-3+ | 0.1% (0 - 2%) |
| CD8+ PD-1(-) TCF1+ TIM-3+ LAG-3+ | 0.0% (0 - 1%) |

**Supplemental Table 7:** Comparison of CD8+ TILs and TIL subpopulations in immunogenic prostate cancer versus metastatic renal cell carcinoma

|  | Immunogenic Prostate Cancer | | Metastatic Renal Cell Carcinoma | |
| --- | --- | --- | --- | --- |
|  | Percentage of CD8+ Cells *Median (Range)* | Density  (cells/mm2) *Median (Range)* | Percentage of CD8+ Cells *Median (Range)* | Density  (cells/mm2) *Median (Range)* |
| CD8+ | - | 253  (39 - 973) | - | 244  (17 - 2907) |
| CD8+ PD-1+ | 42%  (15 - 84%) | 116  (6 - 484) | 59%  (8 - 96%) | 143  (5 - 2674) |
| CD8+ PD-1+  TIM-3(-) LAG-3(-) | 29%  (10 - 60%) | 76  (4 - 460) | 45%  (5 - 89%) | 110  (5 - 1679) |
| CD8+ PD-1+ TCF1+ TIM-3(-) LAG-3(-) | 9%  (0.6 - 23%) | 11  (1 - 204) | 2%  (0 - 43%) | 4  (0 - 317) |

**Supplemental Figure 1**: (A) Analyzer output of TCR sequencing and (B) percentage of productive rearrangements from the top 10 rearrangements (blue) versus all others (grey)

A.

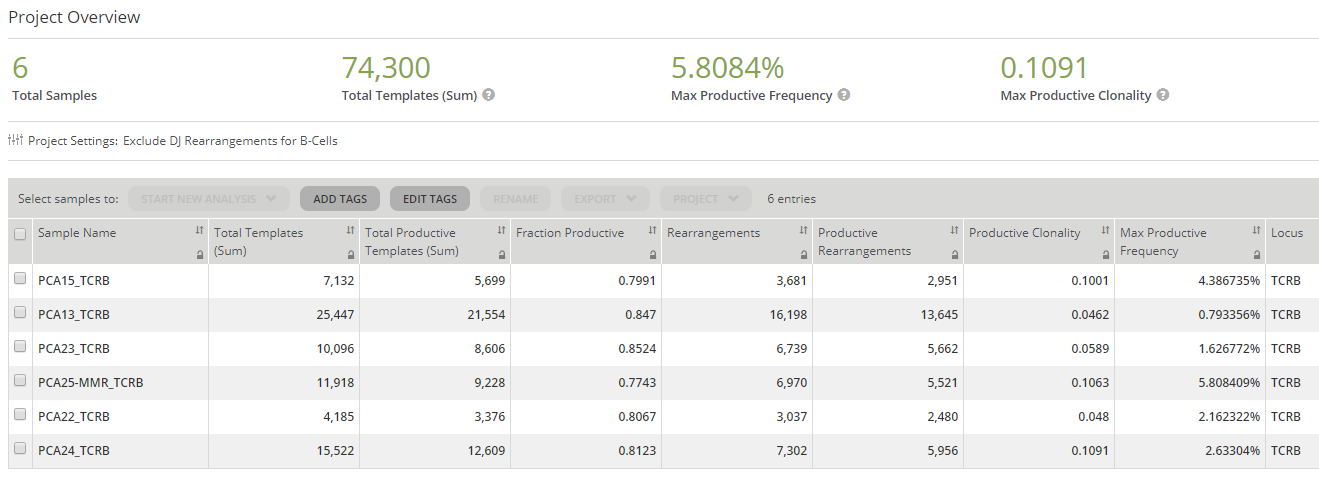

B.

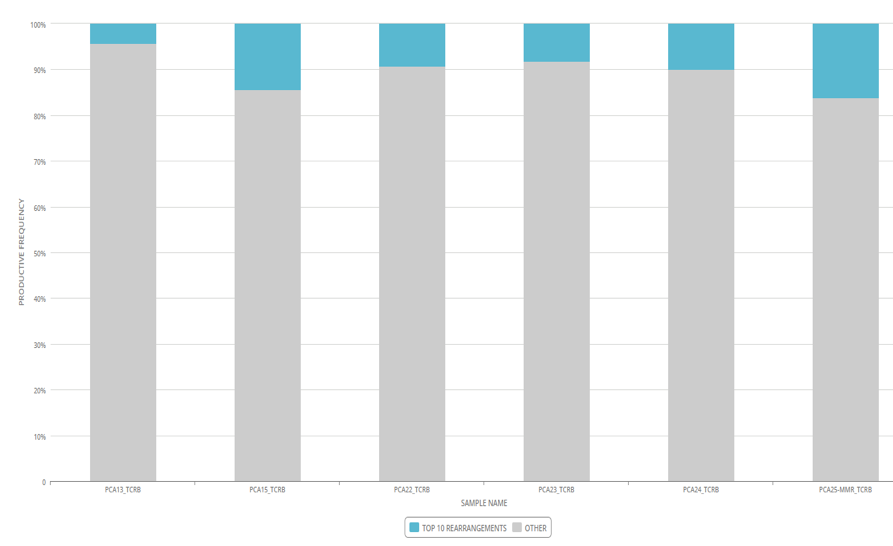

**Supplemental Figure 2:** Copy-number variation in *RB1, BRCA2,* and *CHD1* in individual patients and tumor foci (“cold” indicating non-inflamed focus, “hot” indicating inflamed focus)

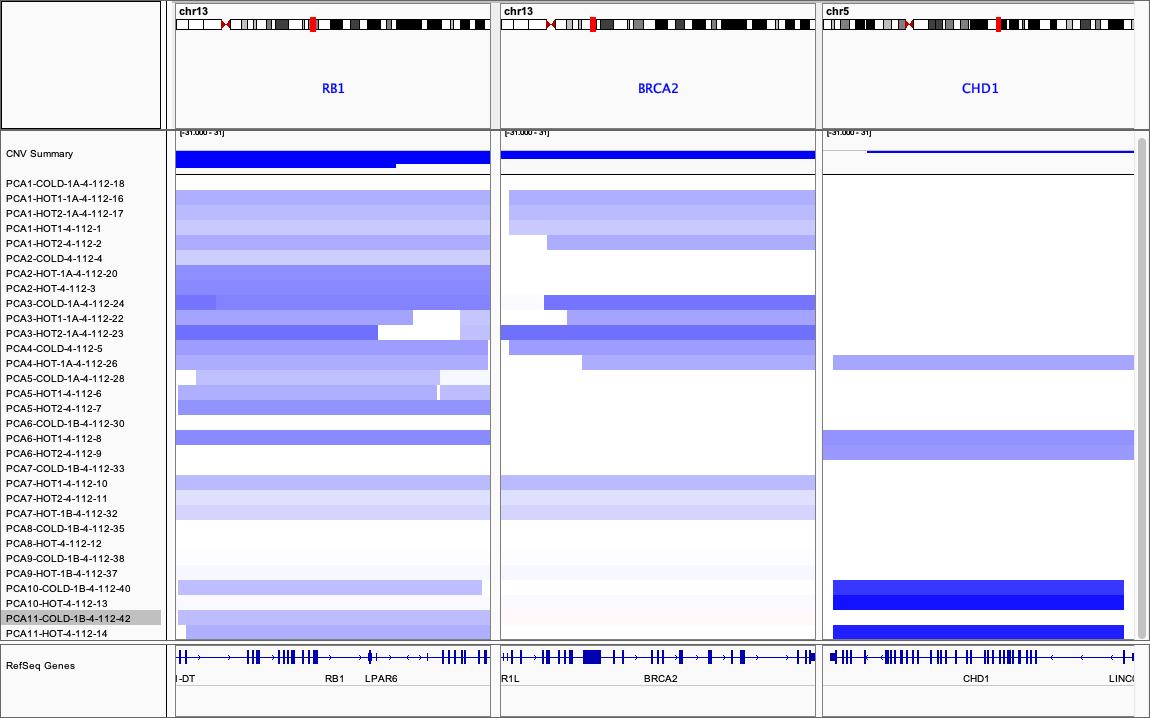

**Supplemental Figure 3:** Complete genomic alterations present in immunogenic and non-immunogenic foci (without first panel showing top alterations, shown in main **Figure 4**)

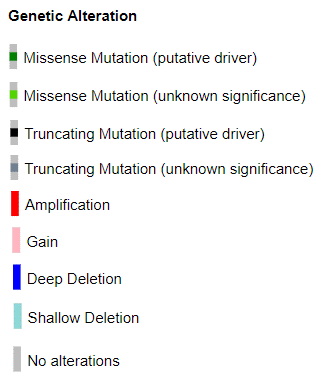

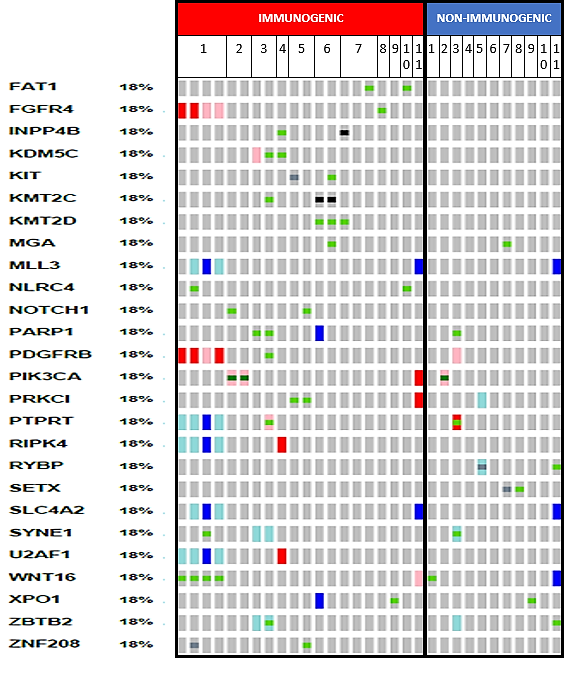

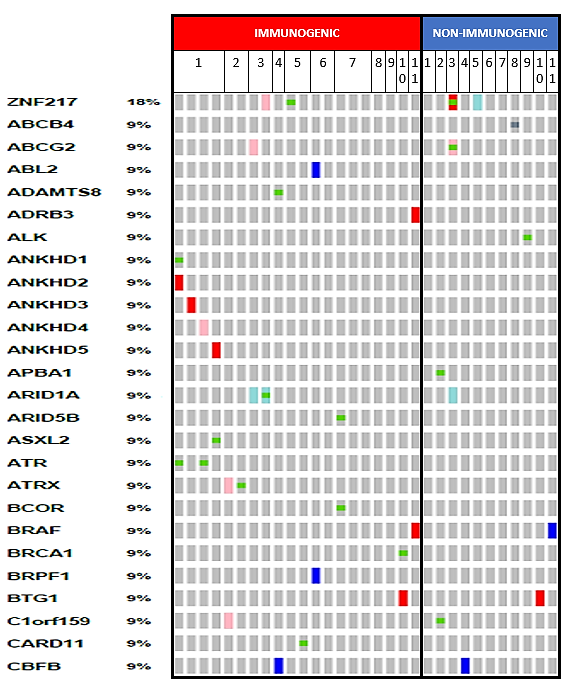

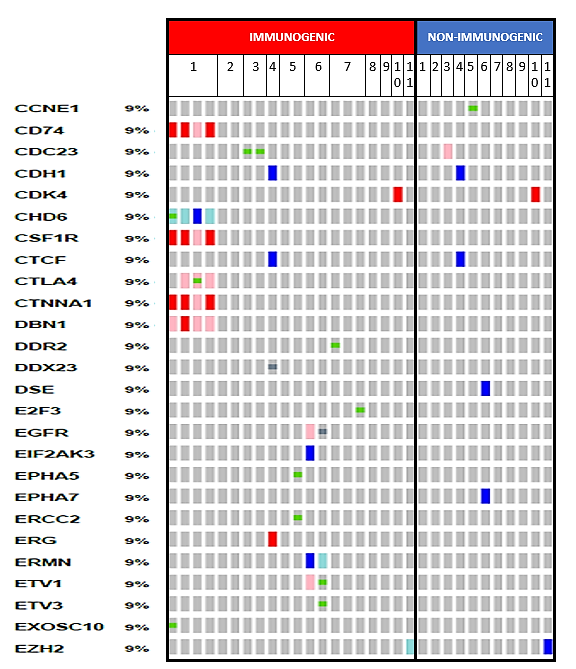

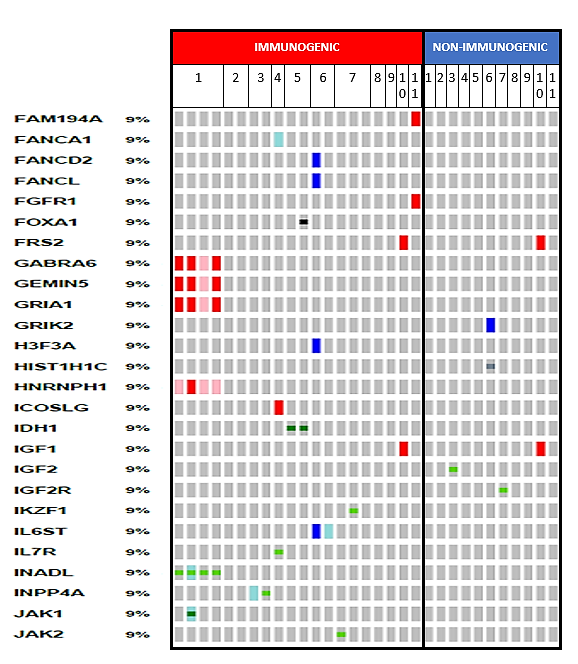

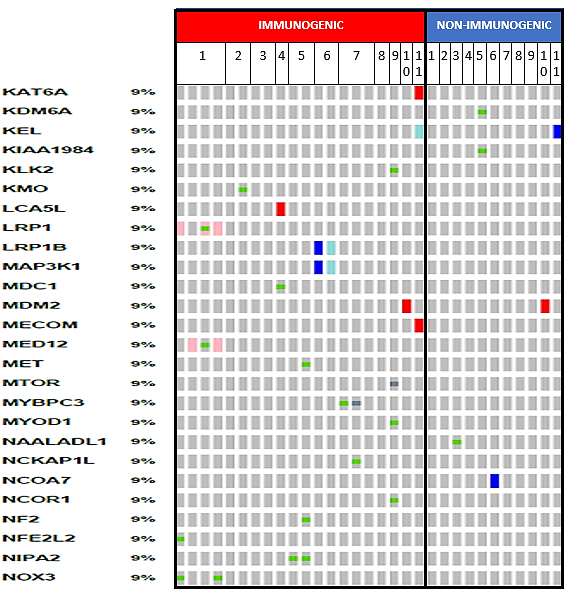

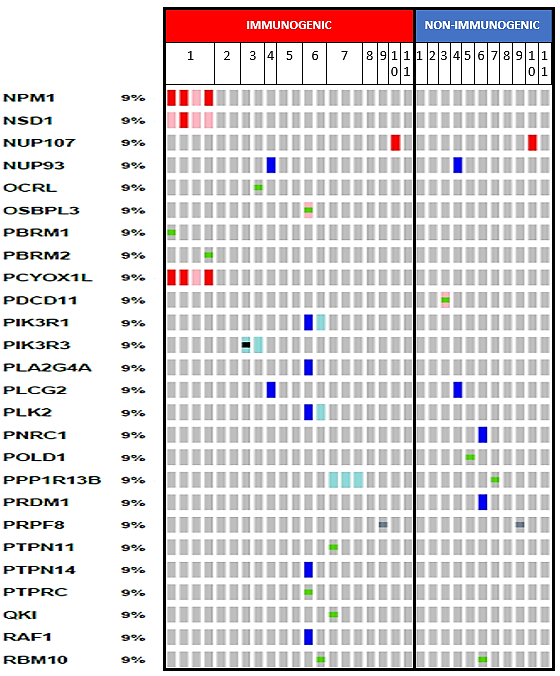

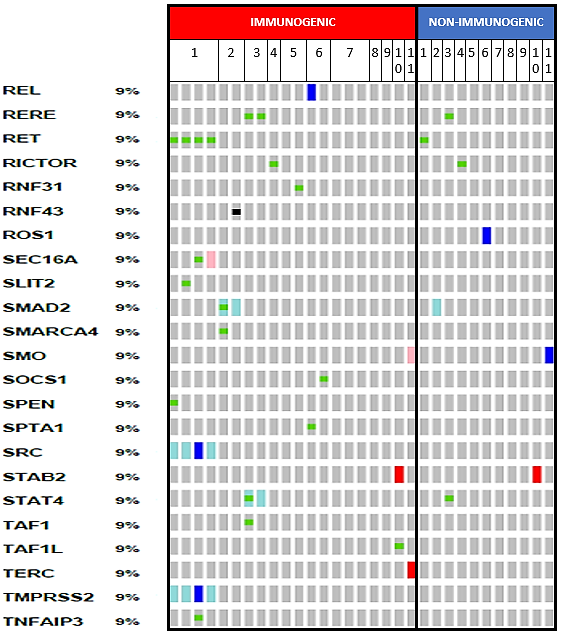

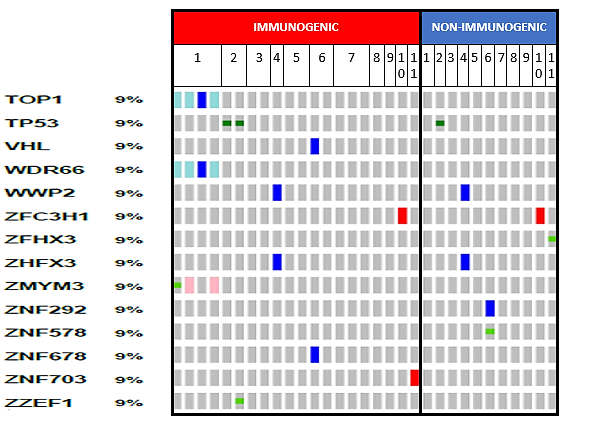

**Supplemental Figure 4:** Most common genomic alterations in immunogenic foci

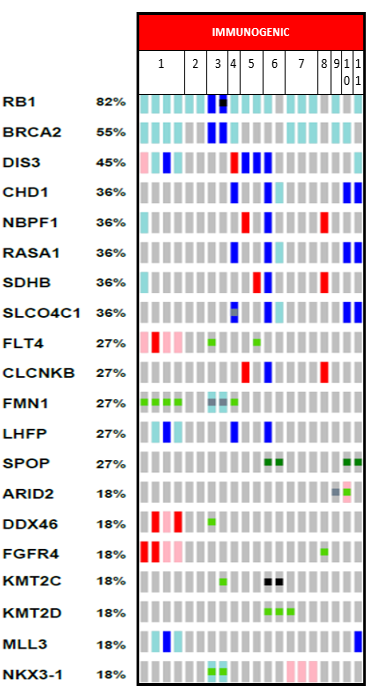

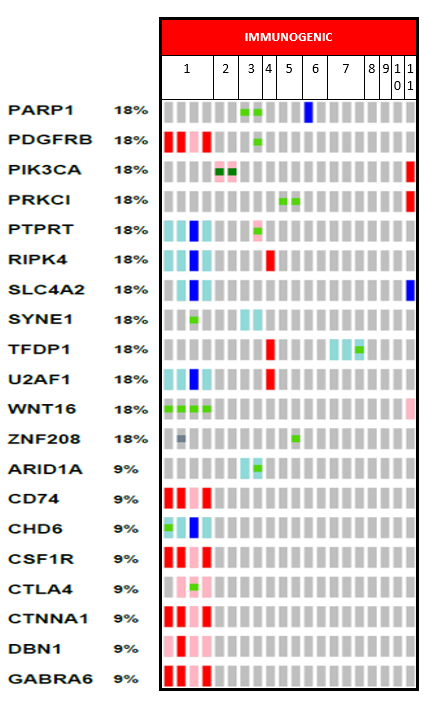

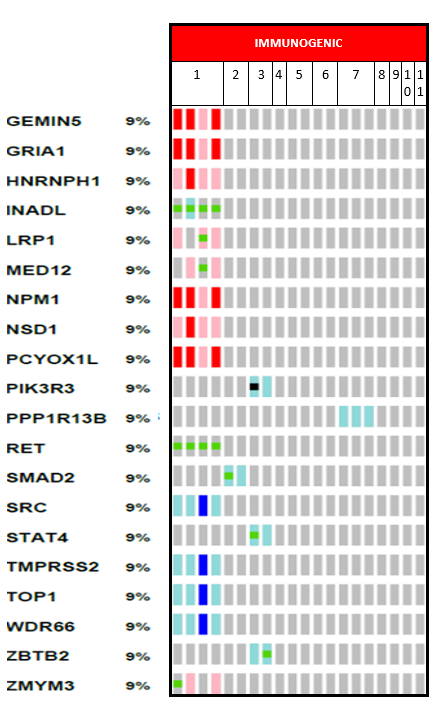

**Supplemental Figure 5:** Most common alterations in non-immunogenic foci

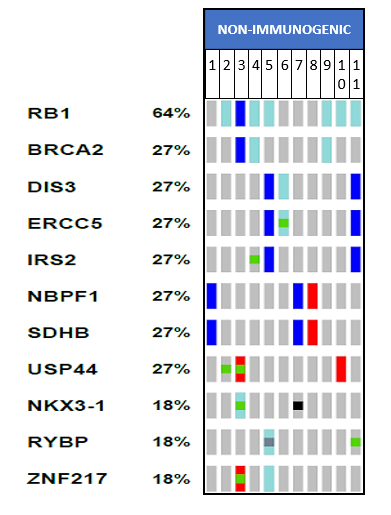
